# Safety of COVID-19 vaccines, their components or their platforms for pregnant women: A rapid review

**DOI:** 10.1101/2021.06.03.21258283

**Authors:** Agustín Ciapponi, Ariel Bardach, Agustina Mazzoni, Tomás Alconada, Steven Anderson, Fernando J. Argento, Jamile Ballivian, Karin Bok, Daniel Comandé, Emily Erbelding, Erin Goucher, Beate Kampmann, Ruth Karron, Flor M. Munoz, María Carolina Palermo, Edward P. K. Parker, Federico Rodriguez Cairoli, María Victoria Santa, Andy Stergachis, Gerald Voss, Xu Xiong, Natalia Zamora, Sabra Zaraa, Mabel Berrueta, Pierre M. Buekens

## Abstract

**Background:** Pregnant women with COVID-19 are at an increased risk of severe COVID-19 illness as well as adverse pregnancy and birth outcomes. Many countries are vaccinating or considering vaccinating pregnant women with limited available data about the safety of this strategy. Early identification of safety concerns of COVID-19 vaccines, including their components, or their technological platforms is therefore urgently needed.

**Methods:** We conducted a rapid systematic review, as the first phase of an ongoing full systematic review, to evaluate the safety of COVID-19 vaccines in pregnant women, including their components, and their technological platforms (whole virus, protein, viral vector or nucleic acid) used in other vaccines, following the Cochrane methods and the PRISMA statement for reporting (PROSPERO-CRD42021234185).

We searched literature databases, COVID-19 and pregnancy registries from inception February 2021 without time or language restriction and explored the reference lists of relevant systematic reviews retrieved. We selected studies of any methodological design that included at least 50 pregnant women or pregnant animals exposed to the vaccines that were selected for review by the COVAX MIWG in August 2020 or their components or platforms included in the COVID-19 vaccines, and evaluated adverse events during pregnancy and the neonatal period.

Pairs of reviewers independently selected studies through the COVIDENCE web software and performed the data extraction through a previously piloted online extraction form. Discrepancies were resolved by consensus.

**Results:** We identified 6768 records, 256 potentially eligible studies were assessed by full-text, and 37 clinical and non-clinical studies (38 reports, involving 2,397,715 pregnant women and 56 pregnant animals) and 12 pregnancy registries were included.

Most studies (89%) were conducted in high-income countries. The most frequent study design was cohort studies (n=21), followed by surveillance studies, randomized controlled trials, and registry analyses. Most studies (76%) allowed comparisons between vaccinated and unvaccinated pregnant women (n=25) or animals (n=3) and reported exposures during the three trimesters of pregnancy.

The most frequent exposure was to AS03 adjuvant in the context of A/H1N1 pandemic influenza vaccines (n=24), followed by aluminum-based adjuvants (n=11). Aluminum phosphate was used in Respiratory Syncytial Virus Fusion candidate vaccines (n=3) and Tdap vaccines (n=3). Different aluminum-based adjuvants were used in hepatitis vaccines. The replication-deficient simian adenovirus ChAdOx1 was used for a Rift Valley fever vaccine. Only one study reported exposure to messenger RNA (mRNA) COVID-19 vaccines that also used lipid nanoparticles. Except for one preliminary report about A/H1N1 influenza vaccination (adjuvant AS03) - corrected by the authors in a more thorough analysis, all studies concluded that there were no safety concerns.

**Conclusion:** This rapid review found no evidence of pregnancy-associated safety concerns of COVID-19 vaccines that were selected for review by the COVAX MIWG or of their components or platforms when used in other vaccines. However, the need for further data on several vaccine platforms and components is warranted given their novelty. Our findings support current WHO guidelines recommending that pregnant women may consider receiving COVID-19 vaccines, particularly if they are at high risk of exposure or have comorbidities that enhance the risk of severe disease.

## BACKGROUND

The COVID-19 Vaccines Global Access Facility (COVAX) is a multilateral initiative to ensure that all countries have fair and equitable access to Coronavirus Disease 2019 (COVID-19) vaccines. Co-led by the GAVI, the Vaccine Alliance, the Coalition for Epidemic Preparedness Innovations (CEPI), and the World Health Organization (WHO), COVAX is a voluntary arrangement that enables countries to pool their resources and risk by collectively investing in vaccine candidates while developing the political and logistical infrastructure needed for vaccine distribution in a transparent and coordinated manner[1-3]. Preauthorization clinical trials of COVID-19 vaccines excluded pregnant women, and only limited human data on their safety during pregnancy was available at the time of emergency use authorization[4]. However, pregnant women with COVID-19 are at increased risk of adverse pregnancy and birth outcomes and severe illness compared to non-pregnant women[5-9]. Many countries are vaccinating or considering vaccinating pregnant women especially if they are at risk of being exposed and even with limited available data about the safety of this strategy. Consequently, it is imperative to identify early safety concerns of COVID-19 vaccines, their components or their platforms; defined as any underlying technology -a mechanism, delivery method, or cell line-that can be used to develop multiple vaccines: whole virus, protein, viral vector, or nucleic acid.

The main characteristics of the vaccines that were selected for review by the COVAX MIWG in August 2020 are presented in **Table 1**. To assist pregnant women, make a more fully informed decision, we aimed to identify safety concerns for pregnant women associated with these exposures through a rapid review of the literature databases as the first phase of an ongoing full systematic review. Given the urgency of the issue for current public health practice across the globe, we performed a rapid review as an interim analysis.

**Table 1.**
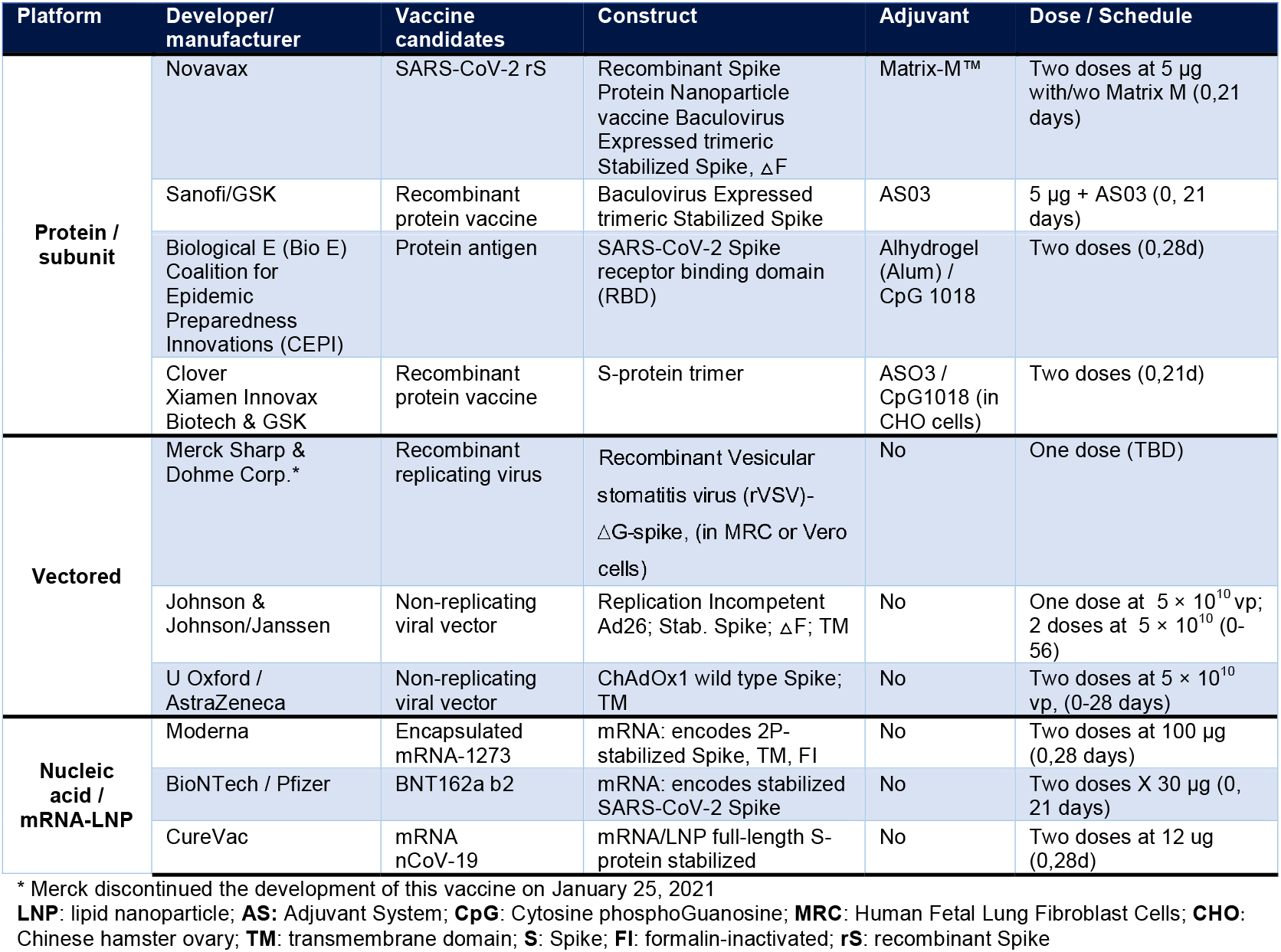
Main characteristics of the vaccines that were selected for review by the COVAX MIWG in August 2020.

## OBJECTIVES

To evaluate the effects of COVID-19 the vaccines that were selected for review by the COVAX MIWG in August 2020, or their components used in other vaccines, on pregnancy safety outcomes.

## METHODS

For this rapid review, we followed the Cochrane methods[10, 11] and the 2020 Preferred reporting items for systematic reviews and meta-analyses (PRISMA) statement[12] for reporting results. This review was registered in PROSPERO (CRD42021234185).

### Inclusion criteria

We included studies that used comparative and non-comparative study designs. Case series were included only if they reported on more than 50 exposed pregnant women. We included also experimental studies of any size with exposed pregnant animals. We excluded systematic reviews (SRs) but explored their reference lists as an additional primary study source.

The interventions or exposures of interest are the COVID-19 candidate vaccines that were selected for review by the COVAX MIWG in August 2020 or vaccine platforms (protein/subunit, vectored, nucleic acid/mRNA-LNP) or components (antigen, vehicle, construct, adjuvants, lipid nanoparticles or other components) used by COVID-19 vaccines. It was considered mandatory that at least one of these exposures was explicitly reported in the report.

We considered outcomes concerning exposure to the vaccines based on the reported gestational age at vaccination (based on validated methods including ultrasound or last menstrual period [LMP] for human studies). We used the 21 standardized case definitions developed by the Global Alignment of Immunization Safety Assessment in Pregnancy (GAIA) of prioritized obstetric and neonatal outcomes based on the Brighton Collaboration process.[13] The ten GAIA obstetric outcomes include hypertensive disorders of pregnancy, maternal death, non-reassuring fetal status, pathways to preterm birth, postpartum hemorrhage, abortion/miscarriage, antenatal bleeding, gestational diabetes, dysfunctional labor, and fetal growth retardation. The 11 neonatal outcomes include congenital anomalies, neonatal death, neonatal infections, preterm birth, stillbirth, low birth weight, small for gestational age, neonatal encephalopathy, respiratory distress, failure to thrive and microcephaly.

Safety outcomes were analyzed according to the US Food and Drug Administration (FDA) Toxicity Grading Scale for Healthy Adult and Adolescent Volunteers Enrolled in Preventive Vaccine Clinical Trials.[14] An adverse event (AE) was defined as any untoward medical occurrence in a patient or clinical investigation subject administered a pharmaceutical product regardless of its causal relationship to the study treatment[15]. An AE can therefore be any unfavorable and unintended sign (including an abnormal laboratory finding), symptom, or disease temporally associated with the use of a medicinal (investigational) product. These include local reactions at the injection site (pain, tenderness, erythema, edema, pruritus, other) and systemic reactions (fever ≥ 38°C or 100.4°F, headache, malaise, myalgia, fatigue, etc.). We also considered other post-vaccination medical events (unsolicited in the studies, reported by organ system as per Medical Dictionary for Regulatory Activities - MedDRA)[16].

We used the following classification to grade the severity of AEs:

⍰ <u>Mild (Grade 1)</u>: Events require minimal or no treatment and do not interfere with the subject’s daily activities.
⍰ <u>Moderate (Grade 2)</u>: Events result in a low level of inconvenience or concern with therapeutic measures. Moderate events may cause some interference with functioning and daily activities.
⍰ <u>Severe (Grade 3)</u>: Events interrupt the subject’s daily activities and may require systemic drug therapy or other treatment. Severe events are usually incapacitating.
⍰ <u>Potentially Life Threatening (Grade 4)</u>: Potentially life-threatening symptoms causing inability to perform basic self-care functions with intervention indicated to prevent permanent impairment, persistent disability, or death.

We also considered other classifications of AEs commonly reported in safety studies, including:

‐ Medically attended adverse events (MAEs): events leading to an otherwise unscheduled visit to or from medical personnel for any reason, including visits to an accident and emergency department)
‐ Serious adverse events (SAEs): resulted in death, were life-threatening, required hospitalization or prolongation of existing hospitalization, resulted in disability/incapacity, or a congenital anomaly/birth defect in the child of a study participant)
‐ Adverse events of special interest (AESIs): events worthy closer follow-up over 6 months post-vaccination. These include vaccine associated enhanced disease such as multisystem inflammatory syndrome (MIS-C/A).

The operative definitions of each specific AEs were reported elsewhere (PROSPERO-CRD42021234185). For this rapid review we considered the integrative outcome “safety concerns” as any statistically significant adverse outcome reported in the comparative studies or unexpected figures, with respect to the published incidences in the peer-reviewed literature, reported in uncontrolled studies.

### Search strategy

We searched published and unpublished studies without restrictions on language or publication status from inception to February 2021 (See **Appendix 1**) in the Cochrane Library databases, MEDLINE, EMBASE, Latin American and Caribbean Health Sciences Literature (LILACS), Science Citation Index Expanded (SCI-EXPANDED), China Network Knowledge Information (CNKI), WHO Database of publications on SARS CoV2, TOXLine, pre-print servers (ArXiv, BiorXiv, medRxiv, search.bioPreprint), and COVID-19 research websites (PregCOV-19LSR, Maternal and Child Health, Nutrition: John Hopkins Centre for Humanitarian health, the LOVE database).

We also searched reference lists of relevant primary studies and systematic reviews retrieved by the search strategy, and adverse events/safety reported in active COVID-19 and pregnancy registries. Additionally, we reviewed information collected in COVID-19 and pregnancy registries. The Food and Drug Administration (FDA), the European Medicines Agency (EMA), clinical trials websites will be searched for the full review. We will then contact original authors and experts in the field for clarification or to obtain extra information. For the full review we will re-run the search strategy to capture any new evidence in databases between March 2021 and current date and time.

### Selection of studies, data extraction and assessment of risk of bias in included studies

Pairs of authors independently screened each identified record by title and abstract and retrieved all the full texts of the potentially eligible studies. Pairs of review authors independently examined the full□text articles for compliance with the inclusion criteria and selected eligible studies. We resolved any disagreements by discussion. We documented the selection process with a ‘PRISMA’ flow chart[12]. This process was conducted through COVIDENCE,[17] a software for systematic reviews.

Pairs of review authors independently extracted data from eligible studies using a data extraction form designed and pilot□tested by the authors. We resolved any disagreements by discussion. Extracted data included study characteristics and outcome data. Where studies have multiple publications, we collated multiple reports of the same study under a single study ID with multiple references.

In **Appendix 2** we describe the risk of bias assessment tools used for each study design. Briefly, we independently independently assessed the risk of bias of the included clinical trials using the Cochrane risk of bias assessment tool[18]. We used the Cochrane EPOC group tools[19] to assess controlled before□after studies (CBAs), nationwide uncontrolled before□after studies (UBAs), interrupted time series (ITSs); Controlled-ITSs (CITSs). We rated the risk of bias in each domain as “low”, “high”, or “unclear”. For observational cohort, case-control, cross-sectional and case-series studies we used the NIH Quality Assessment Tool[20]. After answering the different signaling questions ‘Yes’, ‘No’, ‘Cannot determine’, ‘Not applicable’ or ‘Not reported’ the raters classified the study quality as “good”, “fair”, or “poor”. For consistency with the other designs, we use the classifications low, high or unclear risk of bias respectively.

### Data synthesis

The primary analysis was the comparison of participants exposed and unexposed to the vaccines or their components. Data from non-comparative studies, including numerous registries, were collected and analyzed in the context of the expected global or country incidences of neonatal or obstetric outcomes.

For dichotomous data, we used the numbers of events in the control and intervention groups of each study to calculate Risk Ratios (RRs), Hazard Ratios (HRs) or Mantel□Haenszel Odds Ratios (ORs) or (for very rare events) Peto ORs. For continuous data, we calculated mean difference (MD) or standardized mean difference (SMD) between treatment groups depending on the use of the same or different scales, respectively. We treated large ordinal data as continuous data. We present 95% confidence intervals (CIs) for all outcomes. Where data to calculate RRs/ORs or MDs/SMDs were not available, we utilized the most detailed numerical data available that may facilitate similar analyses of included studies (e.g., test statistics, P values). We assessed whether the estimates calculated in the review for individual studies are compatible in each case with the estimates reported in the study publications. For randomized controlled trials (RCTs), we analyzed the data on an intention□to□treat basis as far as possible (i.e., including all randomized participants in analysis, in the groups to which they were randomized). For this rapid review we tabulated the study intervention/exposition characteristics and compared against the unexposed. We analyze the results of each study to determine any safety concerns as ‘Yes’, ‘No’ or ‘Unclear’.

We planned to conduct subgroup analyses by the trimester of exposure and sensitivity analysis restricted to studies with low risk of bias. However, these were not pursued for this rapid review given the lack of safety concerns identified. We plan to perform meta-analysis and present GRADE ‘Summary of findings’ tables[10, 21] for the full review as was previously stated (PROSPERO-CRD42021234185).

## RESULTS

We identified 6756 records, 265 potentially eligible studies were assessed by full-text and we included 37 clinical and non-clinical studies, involving 2,397,715 pregnant women and 56 pregnant animals from 38 reports[4, 22-58] (**Fig 1**).

**Fig 1.**
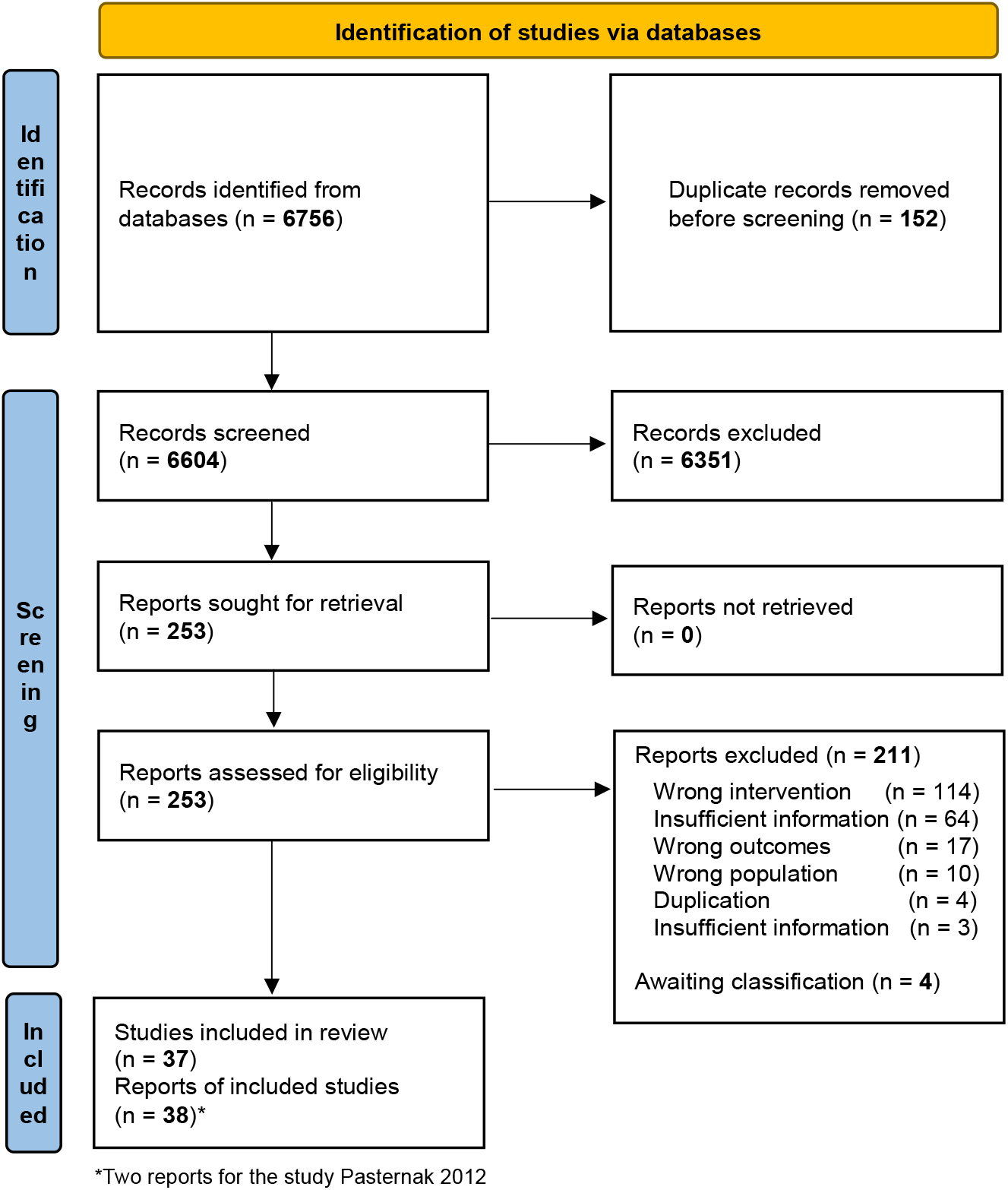
Study flow diagram. (PRISMA 2020)

### Description of studies

The characteristics of included studies are described in **Table 2**. The most frequent study design was cohort studies (n=21) followed by surveillance studies (n=8), controlled trials (n=5) and registry analyses (n=3). Twenty-eight of the included studies (76%) allowed comparisons between vaccinated and unvaccinated pregnant women (n=25) or were conducted in animals (n=3). Nine out of the 37 studies (24%) were abstracts.

**Table 2.**
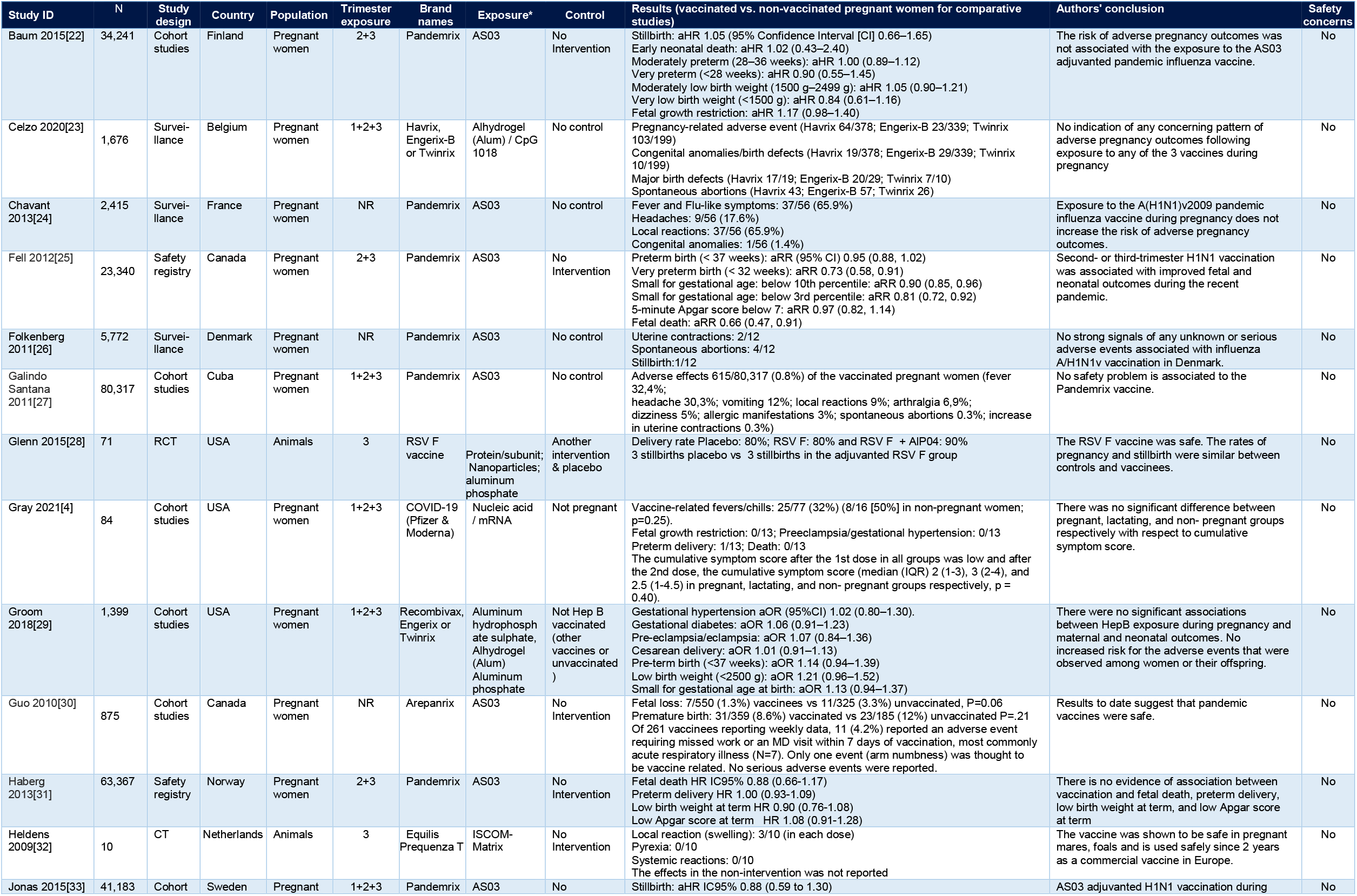
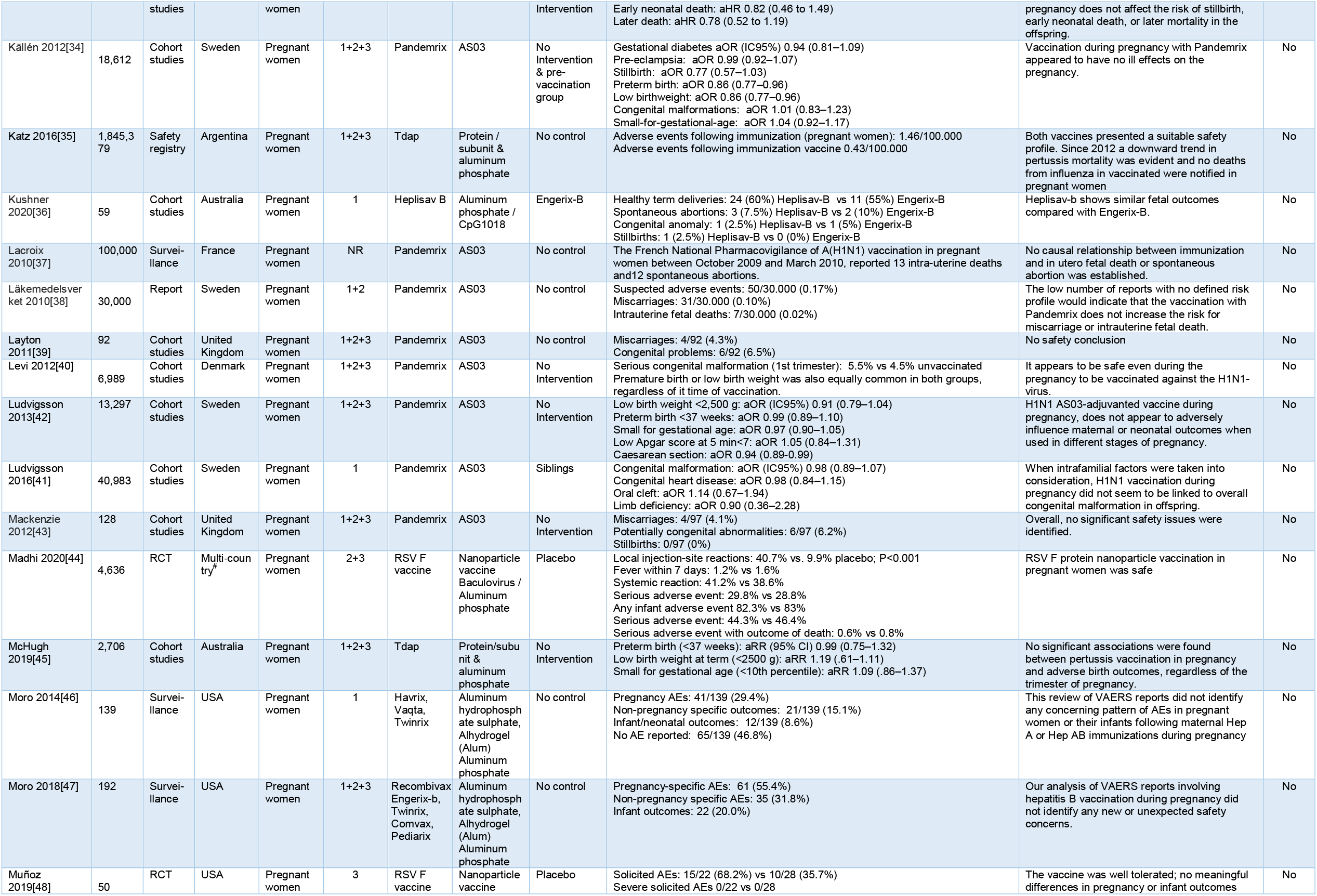
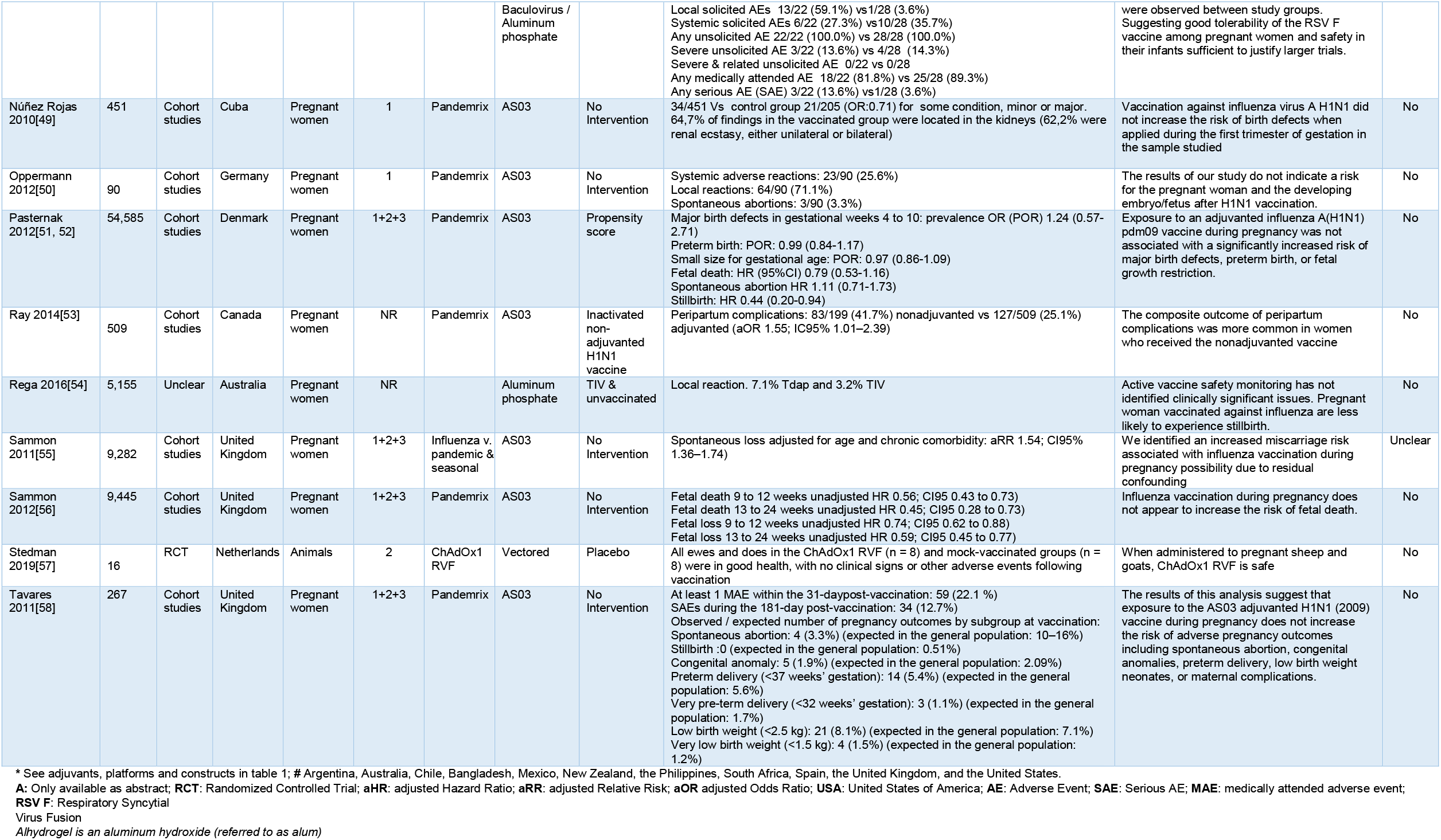
Main characteristics and results of included studies.

The most frequent study location was the USA (n=6), followed by Sweden and United Kingdom (n=5 each), Australia, Canada, and Denmark (n=3 each), Cuba, France and Netherlands (n=2 each), and Argentina, Belgium, Finland, Germany, Norway and multi□country (n=1 each). Only 4 out of 37 studies (11%) involved low-to-middle-income countries (LMICs)[27, 35, 44, 49].

Only 3 out of 37 studies were conducted in animals (8%)[28, 32, 57]. Most of the studies reported exposures during the three trimesters (n=17), only the first trimester (n=5), and the second and third trimester (n=4). In six studies the time of exposure was not reported.

The most frequent exposures were to the AS03 adjuvant (536,240 pregnant participants from 23 studies) and aluminum-based adjuvants (1,861,462 pregnant participants from 11 studies) (**Table 3**). AS03 was the adjuvant of several A/H1N1 pandemic influenza vaccines (Pandemrix® and Arepanrix), while the influenza vaccine Equilis® used ISCOM-Matrix[32]. The aluminum phosphate was used in the testing of candidate Respiratory Syncytial Virus Fusion (RSV F) vaccines in pregnant women [28, 44, 48] (n=3). Aluminum phosphate was also used in Tdap vaccines[35, 45, 54] (n=3). Different aluminum salts were used in Hepatitis vaccines[23, 29, 36, 46, 47]. One study reported on the use of the ChAdOx1 vector for a Rift Valley fever vaccine[57]. Only one study specific to COVID-19 was identified, reporting exposure to mRNA-LNP from Pfizer & Moderna COVID-19 vaccines[4].

**Table 3.** Exposure to vaccine components/platforms.

| Exposure <sup>References</sup> | Pregnant participants | studies (%) | Adjusted relative effects<sup># \$</sup><br>(exposed vs no exposed) |
| --- | --- | --- | --- |
| AS03[22, 24-27, 30, 31, 33, 34, 37-43, 49-53, 55, 56, 58] | 536,240 | 23 (62%) | Congenital malformation: 0.98 to 1.01<br>Fetal death 0.66 to 0.88<br>Early neonatal death: 0.82 to 1.02<br>Later death: 0.78<br>Preterm delivery 0.86 to 1.00 (Very preterm 0.73<sup>\$</sup> to 0.90)<br>Low birth weight/small at term 0.86 to 1.04<br>(<10th & 3rd percentile: 0.90 & 0.81; very low birth weight 0.84)<br>Low Apgar score at term 0.97 to 1.08<br>Gestational diabetes 0.94<br>Pre-eclampsia: 0.99<br>Stillbirth: 0.77 to 1.05<br>Caesarean section: 0.94<br>Peripartum complications: 0.65<sup>\$</sup> |
| *Aluminum phosphate[28, 35, 44, 45, 48] | 1,852,842 | 5 (14%) | Preterm birth: 0.99<br>Low birth weight at term: 1.19<br>Small for gestational age (<10th percentile): 1.09 |
| *Aluminum salts only[29, 46, 47, 54] | 6,885 | 4 (11%) | Stillbirth: 0.49<sup>\$</sup><br>Gestational hypertension 1.02<br>Gestational diabetes: 1.06<br>Pre-eclampsia/eclampsia: 1.07<br>Caesarean delivery: 1.01<br>Pre-term birth (<37 weeks): 1.14<br>Low birth weight (<2500 g): 1.21<br>Small for gestational age at birth: 1.13 |
| *CpG 1018 & Aluminum salts[23, 36] | 1,735 | 2 (5%) | Not available |
| ISCOM-Matrix[32] | 10 | 1 (3%) | Not available |
| mRNA-LNP[4] | 84 | 1 (3%) | Not available |
| ChAdOx1 RVF[57] | 16 | 1 (3%) | Not available |

\* Any aluminum exposure 1,861,462 pregnant participants from 11 studies; LNP: lipid nanoparticle

The 12 COVID-19 and pregnancy registries identified (UKOS, PAN-COVID, BPSU, NPC-19, EPICENTRE, periCOVID, INTERCOVID, PregCOV-19LSR,PRIORITY, COVI-PREG), OTIS/MotherToBaby,CHOPAN, and vsafe registries) are presented in **Table 4**. The list of excluded studies and the reasons for exclusion is presented in **Appendix 3**.

**Table 4.** COVID-19 and Pregnancy Registries.

| Registry | Registered countries | Consent needed for enrollment | Obstetric/pregnancy data |  |  |  |  |  | Neonatal and infant outcome |  |  |  | Samples collected | Study population |  | Registry start date (mm/dd/yyyy) | Vaccination Data |  |  |
| --- | --- | --- | --- | --- | --- | --- | --- | --- | --- | --- | --- | --- | --- | --- | --- | --- | --- | --- | --- |
|  |  |  | SARS-CoV-2 infection | COVID-19 | Early pregnancy loss | Fetal Growth Restriction | Stillbirth | Delivery outcome | Birth condition | Neonatal outcome | Vertical transmission | Infant outcome |  | COVID+ Pregnant w. | COVID- Pregnant w. |  | General population | Pregnant women | Post-vaccination outcomes |
| UKOSS (National) | UK | No | X | X |  | X | X | X | X | X | X | X | No | X |  | 03/01/20 |  | X |  |
| PAN-COVID (International) | 42 countries | Yes | X | X | X | X | X | X | X | X | X |  | No | X | N/A | N/A | N/A | N/A | N/A |
| BPSU (National) | UK | No |  | X |  |  |  | X | X | X | X | X | No |  | N/A | N/A |  |  |  |
| NPC-19 (SONPM/AAP) (National) | USA | No |  | X |  | X | X | X | X | X | X | X | No | X |  | N/A | No | No | No |
| EPICENTRE (International) | 23 European countries<br>Australia | No | X | X |  |  |  | X | X | X | X | X | No | X | N/A | N/A | N/A | N/A | N/A |
| periCOVID (International) | 7 in Africa<br>7 in Europe | Yes | X | X |  | X | X | X | X | X | X | X | Yes* | X |  | N/A | No | No | No |
| INTERCOVID (International) | 29 registered countries (South America & Africa) | Yes | X | X | X | X | X | X | X | X | X | X | No | X | X | 04/20/20 | N/A | N/A | N/A |
| PregCOV-19L SR (International) | Live systematic reviews | No | X | X | X | X | X | X | X | X | X | X | No | X | X | N/A | N/A | N/A | N/A |
| PRIORITY (National) | USA | Yes | X | X | X | X | X | X | X | X | X | X | No | X | X | 03/22/20 | No | No | No |
| COVI-PREG (International) | 23 countries in Asia, Africa, Europe, the Americas and Oceania | Yes | X | X | X | X | X | X | X | X | X | X | No | X | X | N/A | N/A | N/A | N/A |
| OTIS/MotherToBaby (Multi-national) | North America (USA and Canada) | Yes | X | X | X | X | X | X | X | X | X | X | No | X | X | N/A | N/A | N/A | N/A |
| CHOPAN (Multi-national) | Australia, New Zealand | No# | X | X | X | X | X | X | X | X | X | X | No | X | No | N/A |  |  |  |
| vsafe pregnancy registry | USA | Yes | X | X | X | X | X | X | X | X | X | X | No | X | X | N/A |  | X | X |

\*blood, throat/NPA swab, urine, stool, cord blood, placenta, amniotic fluid, breast milk; # Women can opt out

**Table 4.** Risk of bias of clinical trials.

| Study ID | Adequate sequence generation | Allocation concealment | Blinding of participant & personnel | Blinding of outcome assessment | Incomplete outcome data | Selective reporting | Other sources of bias |
| --- | --- | --- | --- | --- | --- | --- | --- |
| Glenn 2015 | Unclear | Unclear | Low | Low | Low | Unclear | Unclear |
| Heldens 2009 | High | High | Unclear | Unclear | Low | Unclear | High |
| Madhi 2020 | High | High | Unclear | Unclear | Low | Unclear | Unclear |
| Muñoz 2019 | Unclear | Unclear | High | Low | Low | Unclear | Unclear |
| Stedman 2019 | Unclear | Unclear | Unclear | Unclear | Low | Unclear | Unclear |

### Risk of bias in included studies

The risk of bias for the included controlled trials are presented in **Table 5** and for the included observational studies in **Table 6**.

**Table 5.** Risk of bias of observational studies.

| Study ID | Study design | Signaling questions* |  |  |  |  |  |  |  |  |  |  |  |  |  | Global Quality |
| --- | --- | --- | --- | --- | --- | --- | --- | --- | --- | --- | --- | --- | --- | --- | --- | --- |
|  |  | 1 | 2 | 3 | 4 | 5 | 6 | 7 | 8 | 9 | 10 | 11 | 12 | 13 | 14 |  |
| Baum 2015 | Cohort studies | Yes | Yes | Yes | Yes | No | Yes | Yes | NA | Yes | No | Yes | No | Yes | Yes | Good |
| Galindo Santana 2011 | Cohort studies | Yes | Yes | Yes | Yes | No | Yes | Yes | NA | Yes | No | Yes | No | NR | No | Fair |
| Gray 2021 | Cohort studies | Yes | Yes | Yes | Yes | No | Yes | Yes | NA | Yes | NA | Yes | NR | Yes | No | Fair |
| Groom 2018 | Cohort studies | Yes | Yes | Yes | Yes | No | Yes | Yes | NA | Yes | NA | Yes | NR | Yes | Yes | Good |
| Guo 2010 | Cohort studies | Yes | CD | CD | CD | CD | Yes | Yes | NA | CD | NA | CD | NR | Yes | No | Fair |
| Jonas 2015 | Cohort studies | Yes | Yes | Yes | Yes | NR | Yes | Yes | NA | Yes | NA | Yes | NR | Yes | Yes | Good |
| Källén 2012 | Cohort studies | Yes | Yes | Yes | Yes | NR | Yes | Yes | NA | Yes | NA | Yes | NR | Yes | NR | Fair |
| Kushner 2020 | Cohort studies | Yes | Yes | Yes | NR | No | Yes | Yes | No | Yes | Yes | Yes | NR | NR | No | Fair |
| Layton 2011 | Cohort studies | Yes | CD | CD | CD | CD | Yes | CD | NA | CD | NA | CD | CD | NR | No | Fair |
| Levi 2012 | Cohort studies | Yes | Yes | CD | CD | CD | Yes | Yes | NA | CD | NA | CD | CD | NR | CD | Fair |
| Ludvigsson 2013 | Cohort studies | Yes | Yes | Yes | Yes | NR | Yes | Yes | NA | Yes | NA | Yes | NR | No | Yes | Good |
| Ludvigsson 2016 | Cohort studies | Yes | Yes | Yes | Yes | No | Yes | Yes | NA | Yes | NA | Yes | NR | Yes | Yes | Good |
| Mackenzie 2012 | Cohort studies | Yes | Yes | Yes | Yes | NR | Yes | Yes | NA | Yes | No | Yes | NR | Yes | No | Fair |
| McHugh 2019 | Cohort studies | Yes | Yes | Yes | Yes | No | Yes | Yes | NA | Yes | NA | Yes | NR | Yes | Yes | Good |
| Núñez Rojas 2010 | Cohort studies | Yes | Yes | NR | No | No | Yes | Yes | NA | Yes | NA | No | NR | NR | No | Poor |
| Oppermann 2012 | Cohort studies | Yes | Yes | Yes | Yes | Yes | Yes | Yes | NA | Yes | NA | Yes | No | Yes | Yes | Good |
| Pasternak 2012 | Cohort studies | Yes | Yes | Yes | Yes | NR | Yes | Yes | NA | Yes | No | Yes | NR | Yes | Yes | Good |
| Pasternak 2012 | Cohort studies | Yes | Yes | Yes | Yes | No | Yes | Yes | NA | Yes | NA | Yes | NA | NA | Yes | Good |
| Ray 2014 | Cohort studies | No | No | CD | Yes | NR | Yes | CD | NA | No | No | No | NR | CD | NR | Poor |
| Sammon 2011 | Cohort studies | Yes | Yes | Yes | Yes | No | Yes | CD | NA | Yes | NA | No | NA | NA | No | Poor |
| Sammon 2012 | Cohort studies | Yes | Yes | Yes | Yes | No | Yes | Yes | NA | Yes | No | Yes | No | NR | Yes | Good |
| Tavares 2011 | Cohort studies | Yes | Yes | CD | Yes | No | Yes | Yes | NA | Yes | No | Yes | No | No | No | Fair |
| Fell 2012 | Registry analysis | Yes | Yes | Yes | Yes | No | Yes | Yes | NA | Yes | NA | Yes | NA | Yes | Yes | Good |
| Haberg 2013 | Registry analysis | Yes | Yes | Yes | Yes | No | Yes | Yes | NA | Yes | NA | Yes | NA | No | Yes | Good |
| Katz 2016 | Registry analysis | Yes | Yes | CD | Yes | No | Yes | CD | NA | Yes | NA | Yes | NA | NA | NA | Fair |
| Celzo 2020 | Surveillance | Yes | Yes | CD | No | No | Yes | NR | NR | Yes | NA | Yes | NA | NA | No | Fair |
| Chavant 2013 | Surveillance | Yes | Yes | NA | Yes | No | Yes | Yes | Yes | Yes | No | Yes | No | CD | No | Poor |
| Folkenberg 2011 | Surveillance | Yes | Yes | CD | Yes | No | Yes | Yes | NA | Yes | NA | Yes | NA | NA | NA | Good |
| Lacroix 2010 | Surveillance | Yes | Yes | CD | Yes | No | No | Yes | NA | Yes | No | Yes | NA | NA | No | Fair |
| Läkemedelsverket 2010 | Surveillance | Yes | Yes | CD | CD | No | Yes | Yes | NA | Yes | NA | No | No | NA | No | Poor |
| Moro 2014 | Surveillance | Yes | Yes | CD | Yes | No | Yes | Yes | NA | Yes | NA | Yes | NA | NA | NA | Good |
| Moro 2018 | Surveillance | Yes | Yes | NA | Yes | No | Yes | Yes | NA | No | No | Yes | NA | NA | NA | Poor |
| Rega 2016 | Surveillance | No | No | Yes | NR | No | Yes | NR | NA | Yes | NA | NR | NA | NA | No | Poor |

NA: not applicable, NR: not reported, CD: cannot be determined
**\*Signaling questions**

We assessed the 38 included reports. Among the five RCTs two (40%) presented high risk of bias in the randomization process and one (20%) in blinding of participant and personnel. Among the 33 observational study reports, 14 were classified as “good” (43%), 12 as “fair” (36%) and 7 as “poor” (21%).

### Outcomes of exposures

The results of included studies are described in **Table 2**. There were 13 pregnancy-related outcomes (26 reports), 8 neonatal outcomes (19 reports) and 9 maternal outcomes (13 reports). The most reported pregnancy outcomes were preterm delivery (n=12), stillbirth (n=9), spontaneous abortion (n=9), fetal growth restriction/small gestational age (n=8), fetal death (n=6). The most reported neonatal outcomes were congenital anomalies (n=9), low birth weight (n=8) and the most reported maternal outcomes were local reactions (n=7), systemic reactions (n=5), serious adverse event (n=6).

The adjusted relative effects comparing exposed vs. no exposed pregnant participants were summarized in **Table 3**. None of the available exposure, including AS03, aluminum phosphate or aluminum salts only, was statistically associated with adverse outcomes. Only AS03 showed a statistically lower frequency of very preterm aRR 0.73 (95%CI 0.58 to 0.91)^[25]^ and peripartum complications aOR 0.65 (95%CI 0.42 to 0.99) [53]; and aluminum salts with lower stillbirth aHR 0.49 (95%CI 0.29 to 0.84)[54]. The lack of more comparative information regarding “safety concerns” precludes further subgroup analysis by exposure.

Of the 37 included studies, 36 (97%) concluded that there was no evidence of safety concerns. Only one study[55], reported as abstract, mentioned unclear safety concerns regarding the 9,026 pregnancies ending in a delivery that had a record of the swine flu vaccine during or just before their pregnancy. The authors reported that they may not have captured early pregnancy losses and that some misclassification of outcome may have occurred or residual confounding after adjusting for age and chronic comorbidity may have been present. In fact the full-text manuscript reported one year later by these authors[56] included 9,445 women vaccinated before or during pregnancy and found no difference in the hazard of fetal loss during weeks 25 to 43 and a lower hazard of fetal loss than unvaccinated pregnancies in gestational weeks 9 to 12 and 13 to 24.

The planned subgroup analyses by the trimester of exposure and sensitivity analysis restricted to studies with low risk of bias were conducted given the lack of reported safety concerns in every study.

**Table 4** shows the characteristics of the 12 identified COVID-19 and pregnancy registries, with potential data on safety/adverse events. The USA and the UK were the most represented countries. Some large registries are multinational, such as EPICENTRE, COVI-PREG or PAN-COVID, which gathers data from 42 countries. Most registries include information on obstetric/pregnancy outcomes like early pregnancy loss, fetal growth, stillbirths, and delivery outcomes. All of them include neonatal and infant outcomes. Additionally, UKOSS and V-safe include specific vaccination information on the pregnant population. PeriCOVID was the only registry that collected blood samples. More detailed information on the relevant information from these registries will be described in the full systematic review, which is currently ongoing.

We also identified three ongoing studies in the COVID-19 vaccine tracker developed by the Vaccine Centre at the London School of Hygiene and Tropical Medicine which contains information of the WHO, the Milken Institute and clinicaltrials.gov. databases[59]. One phase-2 trial is assessing the Ad26.COV2.S vaccine (a monovalent vaccine composed of a recombinant, replication-incompetent adenovirus type 26 vector)[60], and a phase-2/3 trial assessing the BNT162b2 vaccine (an RNA vaccine)[61] are being conducted in the United States, Australia, Brazil, Canada, Finland, South Africa, Spain, and in the United Kingdom. In addition, a phase-4 nonrandomized controlled study is being conducted in Belgium to verify if Sars-Cov-2 specific antibodies can be demonstrated in blood serum and milk of lactating mothers vaccinated with the CX-024414 vaccine (mRNA vaccine)[62].

## DISCUSSION

Through this rapid review, we found no evidence of safety concerns regarding the COVID-19 vaccines that were selected for review by the COVAX MIWG in August 2020, their components or platforms used in other vaccines during pregnancy.

None of the adjusted relative effects comparing exposed vs. no exposed pregnant participants of the available exposure results was statistically associated with adverse outcomes Only AS03 showed a statistically lower frequency of very preterm^[25]^ and peripartum complications [53] and aluminum salts showed lower stillbirth aHR 0.49 (95%CI 0.29 to 0.84)[54]. Uncontrolled studies in general reported low frequencies of adverse outcomes. One study[55], reported as an abstract, suggested safety concerns regarding the swine flu vaccine (AS03 adjuvant) during or just before pregnancy, but the authors recognized potential bias for this finding. The authors published the full-text manuscript[56] one year later and after a more complete analysis they concluded that there is no evidence of safety concerns.

Nine systematic reviews consistently supported the safety of influenza vaccines during pregnancy[63-71]. In general, cohort studies showed the benefits of vaccinating during pregnancy such as significantly decreased risks for preterm birth, small for gestational age and fetal death. However, after adjusting for season at the time of vaccination and countries’ income level, only reduction of fetal death remained significant[67]. There is no evidence of an association between influenza vaccination and serious adverse events in the comparative studies[68]. When assessing only major malformations, no increased risk was detected after immunization at any trimester. Neither adjuvanted nor unadjuvanted vaccines were associated with an increased risk for congenital anomalies[70].

Other systematic reviews also assessed the safety of different vaccines. One SR evaluated the safety of hepatitis B, pneumococcal polysaccharide vaccine and meningococcal polysaccharide vaccines administration during pregnancy and found no clear association with a teratogenic effect on the fetus, preterm labor or spontaneous abortion[72]. Another SR evaluated safety of vaccines frequently given to travelers on pregnant women such as yellow fever, MMR (mumps, measles and rubella), influenza, Tdap (tetanus, diphtheria and pertussis), meningococcus or hepatitis A and B[73]. The authors concluded the safety of influenza vaccine is supported by high-quality evidence. For Tdap vaccine, no evidence of any unexpected harm was found in the meta-analysis of RCTs. Meningococcal vaccines are probably safe during pregnancy, as supported by RCTs comparing meningococcal vaccines to other vaccines. Data supported the safety of hepatitis A, hepatitis B vaccines during pregnancy. In summary primary and secondary evidence described supports the safety of COVID-19 vaccines, their components or their platforms used in other vaccines during pregnancy.

Three recent studies about mRNA-LNP vaccines in pregnant women, published after this rapid review was finalized, reinforced these findings[74-76]. Shimabukuro el al, published preliminary results from the U.S. surveillance review of the safety of mRNA COVID-19 vaccines during pregnancy[75]. The solicited local and systemic reactions reported were similar among persons who identified as pregnant and non-pregnant women. Prabhu et al, studied the antibody response of 122 pregnant women, and their neonates at time of birth, who had received one or both doses of a mRNA-based COVID-19 vaccine[74]. COVID-19 vaccination in pregnancy induced a robust maternal immune response, with transplacental antibody transfer detectable as early as 16 days after the first dose. Rottenstreich et al, 20 pregnant women who received two doses of SARS-CoV-2 BNT162b2 (Pfizer/BioNTech) mRNA vaccine and found similar antibody response[76]. No safety concern was reported in any of these studies. Also, the proportions of adverse pregnancy and neonatal outcomes among completed pregnancies in the registry were similar to the published incidences in pregnant populations studies before the COVID-19 pandemic[77-83].

This rapid review has several strengths. First, we included reports without time, language, or publication type restriction, in human and animals, to provide a timely answer to a hot topic. Second, we adhered to rigorous recommended quality standards to conduct rapid reviews[11] including independent, data extraction and risk of bias assessment and a sensitive and comprehensive search strategy on literature databases to reduce the risk of missing relevant studies. Third, we categorized the exposition to the vaccine components and platforms, which was a challenging issue that demanded frequently to explore additional sources. Finally, we summarized and critically appraised a considerable amount of evidence to conclude if there are safety concerns of the components or platforms used by the vaccines that were selected for review by the COVAX MIWG in August 2020. The vaccines availability has changed over time[84] but we plan to update the search strategy covering the new vaccines for the ongoing full systematic review.

Our study is not exempt from limitations. Only 11% of the total body of evidence comes from LMICs limiting the generalizability to these settings. Additionally, only 76% of included studies allowed comparisons between vaccinated and unvaccinated pregnant women and only five of them were RCTs. Therefore, most of this evidence is observational. Nevertheless, the absence of safety concerns regardless the study design and publication type suggest that this could not be a major limitation. We are aware that the cut-off of 50 animals for non-human studies could be too high, but allowed us to provide timely answers through a rapid review in the context of the pandemics.

Moreover, the set of non-controlled studies do not show unexpected figures with respect to the incidences published in the peer-reviewed literature of neonatal or obstetric outcomes[75]. Regardless of the exposure, all reported a rates of spontaneous abortion in exposed pregnant women described in Table 2 are below the reported the highest global incidence of 31%, or 10% when considering only losses occurring in clinically recognized pregnancies[77]. Tavares 2011 reported a rate of congenital anomalies of 1.9%, in line with the reported rate in the general population of approximately 2 to 4% of livebirths[78-82]. Regarding fetal death, rates reported by Läkemedelsverket 2010 (0.2%) – in Sweden - are consistent with the reported rates of stillbirth for high income countries: approximately 3 deaths per 1000 live births)[85]. None of the included studies conducted in LMICs reported stillbirth rates, which has been reported to be higher than in HIC: approximately 21 deaths per 1000 live births in low income countries[85].

We are aware that the list of Tdap vaccines included our review is incomplete due to the focus of our research question. This vaccine contains aluminum phosphate as adjuvant, which is not used for the COVID-19 vaccines under study, like the alhydrogel adjuvant. Therefore, our search strategy did not include the term “Tdap”. Nevertheless, any aluminum adjuvant retrieved by our search strategy was included and reported.

The nature of this rapid review did not allow us to search in FDA, the EMA websites, clinical trials registers or to contact authors and experts in the field to obtain additional data. For the same reason we could not conduct a meta-analysis that are planned for the full review phase. Regarding COVID-19 and pregnancy registries we identified 12 national or international databases with potentially useful information on safety outcomes. These will be further inspected in the next phase of this work.

Based on existing data, it seems that there are no evident safety risks of COVID-19 vaccines components, or the technological platforms used for pregnant women. It is reasonable to consider COVID-19 vaccination in pregnant women, because their higher risk of adverse outcomes. The next full review phase will add stronger evidence over this important public health issue.

Future experimental data will be needed to assess the pregnancy-related maternal and neonatal COVID-19 vaccine safety. Good quality safety registries, ideally with active surveillance, would also provide an extremely useful evidence from the real-world data.

## Data Availability

All relevant data its presented in the manuscript and supplemental files.

## Acknowledgement

We want to thank Sobanjo-ter Meulen Ajoke, for her supervision and general support and Oduyebo Titilope for her feedback and support.

## Author contributions

All authors contributed to conception and design of this study and AC, FA, AB, MB, SZ, BK, EP, XX, AM and PB prepared the first manuscript draft. All authors interpreted the data, revised critically the first draft, and signed off on the final version.

## Supporting information

Appendix 1. Search strategy

Appendix 2. Risk of bias assessment tools by study design

Appendix 3. List of excluded studies

Appendix 4. PRISMA checklist

## References

[1] Eccleston-Turner M, Upton H. International Collaboration to Ensure Equitable Access to Vaccines for COVID-19: The ACT-Accelerator and the COVAX Facility. Milbank Q. 2021.

[2] Herzog LM, Norheim OF, Emanuel EJ, McCoy MS. Covax must go beyond proportional allocation of covid vaccines to ensure fair and equitable access. BMJ. 2021;372:m4853.

[3] The L. Access to COVID-19 vaccines: looking beyond COVAX. Lancet. 2021;397:941.

[4] Gray KJ, Bordt EA, Atyeo C, Deriso E, Akinwunmi B, Young N, et al. COVID-19 vaccine response in pregnant and lactating women: a cohort study. medRxiv : the preprint server for health sciences. 2021.

[5] Allotey J, Stallings E, Bonet M, Yap M, Chatterjee S, Kew T, et al. Clinical manifestations, risk factors, and maternal and perinatal outcomes of coronavirus disease 2019 in pregnancy: living systematic review and meta-analysis. BMJ (Clinical research ed). 2020;370:m3320.

[6] Figueiro-Filho EA, Yudin M, Farine D. COVID-19 during pregnancy: an overview of maternal characteristics, clinical symptoms, maternal and neonatal outcomes of 10,996 cases described in 15 countries. J Perinat Med. 2020.

[7] Vergara-Merino L, Meza N, Couve-Perez C, Carrasco C, Ortiz-Munoz L, Madrid E, et al. Maternal and perinatal outcomes related to COVID-19 and pregnancy: overview of systematic reviews. Acta Obstet Gynecol Scand. 2021.

[8] Zambrano LD, Ellington S, Strid P, Galang RR, Oduyebo T, Tong VT, et al. Update: Characteristics of Symptomatic Women of Reproductive Age with Laboratory-Confirmed SARS-CoV-2 Infection by Pregnancy Status - United States, January 22-October 3, 2020. MMWR Morb Mortal Wkly Rep. 2020;69:1641–7.

[9] Ciapponi A, Bardach A, Comandé D, Berrueta M, Argento FJ, Cairoli FR, et al. COVID-19 and pregnancy: An umbrella review of clinical presentation, vertical transmission, and maternal and perinatal outcomes. medRxiv. 2021.2021.04.29.21256327.

[10] Higgins J, Thomas J, Chandler J, MS. C, Li T, Page M, et al. Cochrane Handbook for Systematic Reviews of Interventions version 6.0 (updated August 2019). Cochrane, 2019. In: Cochrane, editor.2019.

[11] Garritty C, Gartlehner G, Nussbaumer-Streit B, King VJ, Hamel C, Kamel C, et al. Cochrane Rapid Reviews Methods Group offers evidence-informed guidance to conduct rapid reviews. J Clin Epidemiol. 2020;130:13–22.

[12] Page MJ, McKenzie JE, Bossuyt PM, Boutron I, Hoffmann TC, Mulrow CD, et al. The PRISMA 2020 statement: An updated guideline for reporting systematic reviews. PLoS Med. 2021;18:e1003583.

[13] Kohl KS, Bonhoeffer J, Chen R, Duclos P, Heijbel H, Heininger U, et al. The Brighton Collaboration: enhancing comparability of vaccine safety data. Pharmacoepidemiol Drug Saf. 2003;12:335–40.

[14] CBER-FDA. Guidance for Industry -Toxicity Grading Scale or Healthy Adult and Adolescent-Volunteers Enrolled in Preventive Vaccine Clinical Trials. In: Services USDoHaH, Administration F-FaD, Research C-CfBEa, editors. 2007.

[15] EMA. ICH Topic E 2 A Clinical Safety Data Management: Definitions and Standards for Expedited Reporting EMA - European Medical Agency; 1995.

[16] Fescharek R, Kübler J, Elsasser U, Frank M, Güthlein P. Medical Dictionary for Regulatory Activities (MedDRA). International Journal of Pharmaceutical Medicine. 2004;18:259–69.

[17] Covidence systematic review software. Melbourne, Australia: Veritas Health Innovation.

[18] Higgins J, Green S, (editors). Cochrane Handbook for Systematic Reviews of Interventions Version 5.1.0 [updated March 2011]. In: Cochrane, editor. 2011.

[19] EPOC EPOoCCG. What study designs should be included in an EPOC review? EPOC Resources for review authors, 2017. 2017.

[20] NIH. Study Quality Assessment Tools In: NIH National Heart L, and Blood Institute (NHLBI), editor.2020.

[21] Jan Brozek AO, Holger Schünemann. The GRADE Working Group GRADEpro. 3.2.2 for Windows. Updated March 2009. 2009.

[22] Baum U, Leino T, Gissler M, Kilpi T, Jokinen J. Perinatal survival and health after maternal influenza A(H1N1)pdm09 vaccination: A cohort study of pregnancies stratified by trimester of vaccination. Vaccine. 2015;33:4850–7.

[23] Celzo F, Buyse H, Welby S, Ibrahimi A. Safety evaluation of adverse events following vaccination with Havrix, Engerix-B or Twinrix during pregnancy. Vaccine. 2020;38:6215–23.

[24] Chavant F, Ingrand I, Jonville-Bera AP, Plazanet C, Gras-Champel V, Lagarce L, et al. The PREGVAXGRIP study: a cohort study to assess foetal and neonatal consequences of in utero exposure to vaccination against A(H1N1)v2009 influenza. Drug safety. 2013;36:455–65.

[25] Fell DB, Sprague AE, Liu N, Yasseen AS, 3rd, Wen S-W, Smith G, et al. H1N1 influenza vaccination during pregnancy and fetal and neonatal outcomes. American journal of public health. 2012;102:e33–40.

[26] Folkenberg M, Callreus T, Svanstrom H, Valentiner-Branth P, Hviid A. Spontaneous reporting of adverse events following immunisation against pandemic influenza in Denmark November 2009-March 2010. Vaccine. 2011;29:1180–4.

[27] Galindo Santana BM, Pelaez Sanchez OR, Galindo Sardina MA, Leon Villafuerte M, Concepcion Diaz D, Estruch Rancano L, et al. Active surveillance of adverse effects of Pandemrix vaccine to prevent influenza A(H1N1) in Cuba. Vigilancia activa de eventos adversos a la vacuna Pandemrixpara prevenir la influenza AH1N1 en Cuba. 2011;63:231–8.

[28] Glenn GM, Fries LF, Smith G, Kpamegan E, Lu H, Guebre-Xabier M, et al. Modeling maternal fetal RSV F vaccine induced antibody transfer in guinea pigs. Special Issue: Advancing maternal immunization programs through research in low and medium income countries. 2015;33:6488–92.

[29] Groom HC, Irving SA, Koppolu P, Smith N, Vazquez-Benitez G, Kharbanda EO, et al. Uptake and safety of Hepatitis B vaccination during pregnancy: a Vaccine Safety Datalink study. Vaccine. 2018;36:6111–6.

[30] Guo Y, Allen V, Bujold E, Coleman B, Drews S, Gouin K, et al. Efficacy and safety of pandemic influenza vaccine in pregnancy. Canadian Journal of Infectious Diseases and Medical Microbiology. 2010;21:209.

[31] Haberg SE, Trogstad L, Gunnes N, Wilcox AJ, Gjessing HK, Samuelsen SO, et al. Risk of fetal death after pandemic influenza virus infection or vaccination. The New England journal of medicine. 2013;368:333–40.

[32] Heldens JGM, Pouwels HGW, Derks CGG, Van de Zande SMA, Hoeijmakers MJH. The first safe inactivated equine influenza vaccine formulation adjuvanted with ISCOM-Matrix that closes the immunity gap. Vaccine. 2009;27:5530–7.

[33] Jonas F, Peter S, Cecilia L, Sven C, Anders E, Örtqvist Å, et al. Maternal vaccination against H1N1 influenza and offspring mortality: Population based cohort study and sibling design. BMJ (Online). 2015;351.

[34] Källén B, Olausson PO. Vaccination against H1N1 influenza with Pandemrix® during pregnancy and delivery outcome: A Swedish register study. BJOG: An International Journal of Obstetrics and Gynaecology. 2012;119:1583–90.

[35] Katz N, Neyro S, Carrega MEP, Del Valle Juarez M, Rancaño C, Pasinovich M, et al. Maternal immunization in argentina: The importance of a safety profile analysis. Open Forum Infectious Diseases. 2016;3.

[36] Kushner T, Youhanna J, Walker R, Erby K, Janssen RS. Safety and immunogenicity of Heplisav-B in pregnancy. Hepatology. 2020;72:469A–70A.

[37] Lacroix I, Damase-Michel C, Kreft-Jais C, Castot AC, Montastruc JL. H1N1 influenza vaccine in pregnant women: French pharmacovigilance survey. Drug Safety. 2010;33:908–9.

[38] Läkemedelsverket. Läkemedelsverket. Final summary of adverse drug reaction reports in Sweden with Pandemrix through October 2009-mid April 2010. June 2, 2010. Accessed 23 May 2011 from www.lakemedelsverket.se. 2010.

[39] Layton D, Dryburgh M, MacDonald TM, Shakir SA, MacKenzie IS. Pilot swine flu vaccination active surveillance study: Final results. Drug Safety. 2011;34:889–90.

[40] Levi M. Vaccination against influenza A(H1N1) virus is also safe during pregnancy. Nederlands Tijdschrift voor Geneeskunde. 2012;156.

[41] Ludvigsson JF, Strom P, Lundholm C, Cnattingius S, Ekbom A, Ortqvist A, et al. Risk for congenital malformation with H1N1 influenza vaccine: a cohort study with sibling analysis. Annals of Internal Medicine. 2016;165:848–55.

[42] Ludvigsson JF, Zugna D, Cnattingius S, Richiardi L, Ekbom A, Ortqvist A, et al. Influenza H1N1 vaccination and adverse pregnancy outcome. European Journal of Epidemiology. 2013;28:579–88.

[43] Mackenzie IS, MacDonald TM, Shakir S, Dryburgh M, Mantay BJ, McDonnell P, et al. Influenza H1N1 (swine flu) vaccination: a safety surveillance feasibility study using self-reporting of serious adverse events and pregnancy outcomes. British Journal of Clinical Pharmacology. 2012;73:801–11.

[44] Madhi SA, Polack FP, Piedra PA, Munoz FM, Trenholme AA, Simoes EA, et al. Vaccination of pregnant women with respiratory syncytial virus vaccine and protection of their infants. New England journal of medicine. 2020;383:426–39.

[45] McHugh L, Marshall HS, Perrett KP, Nolan T, Wood N, Lambert SB, et al. The safety of influenza and pertussis vaccination in pregnancy in a cohort of Australian mother-infant pairs, 2012-2015: the FluMum study. Clinical Infectious Diseases. 2019;68:402–8.

[46] Moro PL, Museru OI, Niu M, Lewis P, Broder K. Reports to the vaccine adverse event reporting system after hepatitis a and hepatitis AB vaccines in pregnant women. American Journal of Obstetrics and Gynecology. 2014;210:561.e1-.e6.

[47] Moro PL, Zheteyeva Y, Barash F, Lewis P, Cano M. Assessing the safety of hepatitis B vaccination during pregnancy in the Vaccine Adverse Event Reporting System (VAERS), 1990-2016. Vaccine. 2018;36:50–4.

[48] Munoz FM, Swamy GK, Hickman SP, Agrawal S, Piedra PA, Glenn GM, et al. Safety and Immunogenicity of a Respiratory Syncytial Virus Fusion (F) Protein Nanoparticle Vaccine in Healthy Third-Trimester Pregnant Women and Their Infants. J Infect Dis. 2019;220:1802–15.

[49] Núñez Rojas Y, Orive Rodríguez N, Varona De La Peña F, Bermúdez Velásquez Y, Raad López AF, Muñoz Martínez Y, et al. Vaccination against influenza a H1N1 and the risk of birth defects. VacciMonitor. 2010;19:209.

[50] Oppermann M, Fritzsche J, Weber-Schoendorfer C, Keller-Stanislawski B, Allignol A, Meister R, et al. A(H1N1)v2009: a controlled observational prospective cohort study on vaccine safety in pregnancy. Vaccine. 2012;30:4445–52.

[51] Pasternak B, Svanstrom H, Molgaard-Nielsen D, Krause TG, Emborg HD, Melbye M, et al. Vaccination against pandemic A/H1N1 2009 influenza in pregnancy and risk of fetal death: cohort study in Denmark. BMJ. 2012;344.

[52] Pasternak B, Svanstrom H, Molgaard-Nielsen D, Krause TG, Emborg HD, Melbye M, et al. Risk of adverse fetal outcomes following administration of a pandemic influenza A(H1N1) vaccine during pregnancy. JAMA, Journal of the American Medical Association. 2012;308:165–74.

[53] Ray JG, McGeer AJ, Blake JM, Lebovic G, Smith GN, Yudin MH. Peripartum outcomes: non-adjuvanted v. adjuvanted H1N1 vaccination. CMAJ : Canadian Medical Association journal = journal de l’Association medicale canadienne. 2014;186:137.

[54] Rega A, Moore H, De Klerk N, Effler P. Maternal vaccinations in Australia-uptake, safety and impact. Australian and New Zealand Journal of Obstetrics and Gynaecology. 2016;56:13–4.

[55] Sammon CJ, McGrogan A, Snowball J, De Vries CS. Swine flu vaccination in pregnancy and associated miscarriage risk. Pharmacoepidemiology and Drug Safety. 2011;20:S58–S9.

[56] Sammon CJ, Snowball J, McGrogan A, de Vries CS. Evaluating the Hazard of Foetal Death following H1N1 Influenza Vaccination; A Population Based Cohort Study in the UK GPRD. PLoS ONE. 2012;7.

[57] Stedman A, Wright D, Schreur PJW, Clark MHA, Hill AVS, Gilbert SC, et al. Safety and efficacy of ChAdOx1 RVF vaccine against Rift Valley fever in pregnant sheep and goats. npj Vaccines. 2019;4.

[58] Tavares F, Nazareth I, Monegal JS, Kolte I, Verstraeten T, Bauchau V. Pregnancy and safety outcomes in women vaccinated with an AS03-adjuvanted split virion H1N1 (2009) pandemic influenza vaccine during pregnancy: a prospective cohort study. Vaccine. 2011;29:6358–65.

[59] COVID-19 vaccine tracker. In: https://vac-lshtm.shinyapps.io/ncov_vaccine_landscape/. Accesed May 20th 2021. Vaccine Centre at the London School of Hygiene & Tropical Medicine; 2021.

[60] A Study of Ad26.COV2.S in Healthy Pregnant Participants (COVID-19). https://ClinicalTrials.gov/show/NCT04765384; 2021.

[61] Study to Evaluate the Safety, Tolerability, and Immunogenicity of SARS CoV-2 RNA Vaccine Candidate (BNT162b2) Against COVID-19 in Healthy Pregnant Women 18 Years of Age and Older. https://ClinicalTrials.gov/show/NCT04754594; 2021.

[62] COVID-19: study to detect transfer of SARS-Cov-2 antibodies in breastmilk. https://www.clinicaltrialsregister.eu/ctr-search/trial/2021-000893-27/BE; 2021.

[63] Bratton KN, Wardle MT, Orenstein WA, Omer SB. Maternal influenza immunization and birth outcomes of stillbirth and spontaneous abortion: a systematic review and meta-analysis. Clin Infect Dis. 2015;60:e11–9.

[64] Demicheli V, Jefferson T, Ferroni E, Rivetti A, Di Pietrantonj C. Vaccines for preventing influenza in healthy adults. Cochrane Database Syst Rev. 2018;2:CD001269.

[65] Fell DB, Platt RW, Lanes A, Wilson K, Kaufman JS, Basso O, et al. Fetal death and preterm birth associated with maternal influenza vaccination: systematic review. BJOG. 2015;122:17–26.

[66] Giles ML, Krishnaswamy S, Macartney K, Cheng A. The safety of inactivated influenza vaccines in pregnancy for birth outcomes: a systematic review. Hum Vaccin Immunother. 2019;15:687–99.

[67] Jeong S, Jang EJ, Jo J, Jang S. Effects of maternal influenza vaccination on adverse birth outcomes: A systematic review and Bayesian meta-analysis. PLoS One. 2019;14:e0220910.

[68] Michiels B, Govaerts F, Remmen R, Vermeire E, Coenen S. A systematic review of the evidence on the effectiveness and risks of inactivated influenza vaccines in different target groups. Vaccine. 2011;29:9159–70.

[69] Nunes MC, Aqil AR, Omer SB, Madhi SA. The Effects of Influenza Vaccination during Pregnancy on Birth Outcomes: A Systematic Review and Meta-Analysis. Am J Perinatol. 2016;33:1104–14.

[70] Polyzos KA, Konstantelias AA, Pitsa CE, Falagas ME. Maternal Influenza Vaccination and Risk for Congenital Malformations: A Systematic Review and Meta-analysis. Obstet Gynecol. 2015;126:1075–84.

[71] Zhang C, Wang X, Liu D, Zhang L, Sun X. A systematic review and meta-analysis of fetal outcomes following the administration of influenza A/H1N1 vaccination during pregnancy. Int J Gynaecol Obstet. 2018;141:141–50.

[72] Makris MC, Polyzos KA, Mavros MN, Athanasiou S, Rafailidis PI, Falagas ME. Safety of hepatitis B, pneumococcal polysaccharide and meningococcal polysaccharide vaccines in pregnancy: a systematic review. Drug Saf. 2012;35:1–14.

[73] Nasser R, Rakedzon S, Dickstein Y, Mousa A, Solt I, Peterisel N, et al. Are all vaccines safe for the pregnant traveller? A systematic review and meta-analysis. J Travel Med. 2020;27.

[74] Prabhu M, Murphy EA, Sukhu AC, Yee J, Singh S, Eng D, et al. Antibody Response to Coronavirus Disease 2019 (COVID-19) Messenger RNA Vaccination in Pregnant Women and Transplacental Passage Into Cord Blood. Obstet Gynecol. 2021.

[75] Shimabukuro TT, Kim SY, Myers TR, Moro PL, Oduyebo T, Panagiotakopoulos L, et al. Preliminary Findings of mRNA Covid-19 Vaccine Safety in Pregnant Persons. N Engl J Med. 2021.

[76] Rottenstreich A, Zarbiv G, Oiknine-Djian E, Zigron R, Wolf DG, Porat S. Efficient maternofetal transplacental transfer of anti-SARS-CoV-2 spike antibodies after antenatal SARS-CoV-2 BNT162b2 mRNA vaccination. Clin Infect Dis. 2021.

[77] Magnus MC, Wilcox AJ, Morken NH, Weinberg CR, Haberg SE. Role of maternal age and pregnancy history in risk of miscarriage: prospective register based study. BMJ. 2019;364:869.

[78] Marden PM, Smith DW, McDonald MJ. CONGENITAL ANOMALIES IN THE NEWBORN INFANT, INCLUDING MINOR VARIATIONS. A STUDY OF 4,412 BABIES BY SURFACE EXAMINATION FOR ANOMALIES AND BUCCAL SMEAR FOR SEX CHROMATIN. J Pediatr. 1964;64:357–71.

[79] Leppig KA, Werler MM, Cann CI, Cook CA, Holmes LB. Predictive value of minor anomalies. I. Association with major malformations. J Pediatr. 1987;110:531–7.

[80] Holmes LB. Current concepts in genetics. Congenital malformations. N Engl J Med. 1976;295:204–7.

[81] Mai CT, Isenburg JL, Canfield MA, Meyer RE, Correa A, Alverson CJ, et al. National population-based estimates for major birth defects, 2010-2014. Birth Defects Res. 2019;111:1420–35.

[82] Feldkamp ML, Carey JC, Byrne JLB, Krikov S, Botto LD. Etiology and clinical presentation of birth defects: population based study. BMJ. 2017;357:j2249.

[83] GBD. 2015.Child Mortality Collaborators. Global, regional, national, and selectedsubnational levels of stillbirths, neonatal, infant, and under-5 mortality, 1980-2015: asystematic analysis for the Global Burden of Disease Study 2015. Lancet 2016; 388:1725. 2015.

[84] WHO. Status of COVID-19 Vaccines within WHO EUL/PQ evaluation process. Accessed 12 May 2011 from: https://extranet.who.int/pqweb/sites/default/files/documents/Status_COVID_VAX_04May2021.pdf. 2021.

[85] Wang H, Bhutta ZA, Coates MM, Coggeshall M, Dandona L, Diallo K, et al. Global, regional, national, and selected subnational levels of stillbirths, neonatal, infant, and under-5 mortality, 1980&#x2013;2015: a systematic analysis for the Global Burden of Disease Study 2015. The Lancet. 2016;388:1725–74.

