## Appendix 1 to 4 for "Safety of COVID-19 vaccines, their components or their platforms for pregnant women: A rapid review"

**Appendix 1. Search strategy**

**Structure description in PubMed**

(((**Pregnancy[Mesh]** OR Pregnan*[tiab] OR **Pregnancy Complications[Mesh]** OR **Abortion, Spontaneous[Mesh**] OR Abortion*[tiab] OR Miscarriage*[tiab] OR Gestational[tiab] OR **Parturition[Mesh]** OR Childbirth*[tiab] OR Parturition*[tiab] OR Partum[tiab] OR **Fetus[Mesh]** OR Fetal[tiab] OR Fetus[tiab] OR Maternofetal[tiab] OR Materno Fetal[tiab] OR Fetomaternal[tiab] OR DART[tiab]) **AND** (**Adjuvants, Immunologic[Mesh]** OR Immunoadjuvant*[tiab] OR Immunologic Adjuvant*[tiab] OR Immunological Adjuvant*[tiab] OR Matrix-M*[all] OR Alhydrogel[all] OR Aluminum*[all] OR AS03[all] OR MF59[all] OR CpG 1018[all] OR Recombinant Spike-Protein[all] OR Baculovirus Expressed[all] OR Baculo*[tiab] OR Stabilized Spike[all] OR S-Protein Stabilized[all] OR S-Protein*[all] OR Molecular Clamp[tiab] OR Full-Length[all] OR Replication Incompetent[all] OR Ad26*[all] OR Adenovirus 26[all] OR Ad5*[all] OR Adenovirus 5[all] OR ChAdOx1[all] OR Measles-vector[all] OR V591[all] OR AZD1222[all] OR BNT162b2[all] OR mRNA-1273[all] OR mRNA-LNP[all] OR Vector Expressing[all] OR Influenza Vaccines[Mesh] OR Influenza Vaccine*[tiab] OR Flu Vaccine*[tiab] OR **Saponins[Mesh]** OR Saponin*[tiab] OR **Nanoparticles[Mesh]** OR Nanoparticle*[tiab] OR LNP[tiab] OR Nanocrystal*[tiab] OR Tween 80[all] OR Span 85[all] OR CpG*[all] OR Water Emulsion[tiab] OR HEK293[tiab] OR Arepanrix[all] OR Pandemrix[all] OR FLUAD[all] OR Dynavax[all] OR Hepatitis B Vaccines[Mesh] OR Hepatitis-B Vaccin*[tiab] OR HEPLISAV-B[all] OR Polyethylene Glycol[all]) **AND** (**Vaccination[Mesh] OR Vaccines[Mesh]** OR Vaccin*[tiab]))) **OR** **((Nanoparticles[Mesh]** OR Nanoparticle*[tiab] OR Nanocrystal*[tiab]) AND (Pregnancy[Mesh] OR Pregnan*[tiab] OR DART[tiab])) NOT (Human[Mesh] NOT Animals[Mesh])

**Ovid MEDLINE**

1	exp Pregnancy

2	Pregnan*.ti,ab.

3	exp Pregnancy Complications

4	exp Abortion, Spontaneous

5	Abortion*.ti,ab.

6	Miscarriage*.ti,ab.

7	Gestational.ti,ab.

8	exp Parturition/

9	Childbirth*.ti,ab.

10	Parturition*.ti,ab.

11	Partum.ti,ab.

12	exp Fetus/

13	Fetal.ti,ab.

14	Fetus.ti,ab.

15	Maternofetal.ti,ab.

16	(Materno adj1 Fetal).ti,ab.

17	Fetomaternal.ti,ab.

18	DART.ti,ab.

19	or/1-18

20	exp Adjuvants, Immunologic

21	Immunoadjuvant*.ti,ab.

22	(Immunologic* adj1 Adjuvant*).ti,ab.

23	Matrix-M*.mp.

24	Alhydrogel.mp.

25	Aluminum*.mp.

26	AS03.mp.

27	MF59.mp.

28	CpG1.mp.

29	(Recombinant adj1 Spike-Protein).mp.

30	Baculo*.mp.

31	(Stabilized adj1 Spike).mp.

32	S-Protein*.mp.

33	(Molecular adj1 Clamp).mp.

34	(Full adj1 Length).mp.

35	(Replication adj1 Incompetent).mp.

36	Ad26*.mp.

37	Adenovirus-26.mp.

38	Ad5*.mp.

39	Adenovirus-5.mp.

40	ChAdOx1.mp.

41	Measles-Vector.mp.

42	V591.mp.

43	AZD1222.mp.

44	BNT162b2.mp.

45	mRNA*.mp.

46	(Vector adj1 Expressing).mp.

47	exp Influenza Vaccines/

48	(Influenza adj1 Vaccine*).mp.

49	(Flu adj1 Vaccine*).mp.

50	exp Saponins/

51	Saponin*.mp.

52	exp Nanoparticles/

53	Nanoparticle*.mp.

54	LNP.mp.

55	Nanocrystal*.mp.

56	Arepanrix.mp.

57	Pandemrix.mp.

58	FLUAD.mp.

59	Dynavax.mp.

60	exp Hepatitis B Vaccines/

61	(Hepatitis-B adj1 Vaccin*).mp.

62	HEPLISAV-B.mp.

63	(Polyethylene adj1 Glycol).mp.

64	or/20-63

65	exp Vaccination/

66	exp Vaccines/

67	Vaccin*.ti,ab.

68	or/65-67

69	19 and 64 and 68

70	exp Nanoparticles/

71	Nanoparticle*.mp.

72	LNP.mp.

73	Nanocrystal*.mp.

74	or/70-73

75	exp Pregnancy/

76	Pregnan*.ti,ab.

77	DART.ti,ab.

78	or/75-77

79	exp Animals/

80	74 and 78 and 79

81	69 or 80

**EMBase**

#83. #70 OR #82

#82. #81 NOT #80

#81. #75 AND #79

#80. 'animal'/exp

#79. #76 OR #77 OR #78

#78. dart:ti,ab

#77. pregnan*:ti,ab

#76. 'pregnancy'/exp

#75. #71 OR #72 OR #73 OR #74

#74. nanocrystal*:ti,ab

#73. lnp:ti,ab

#72. nanoparticle*:ti,ab

#71. 'nanoparticle'/exp

#70. #19 AND #65 AND #69

#69. #66 OR #67 OR #68

#68. vaccin*:ti,ab

#67. 'vaccine'/exp

#66. 'vaccination'/exp

#65. #20 OR #21 OR #22 OR #23 OR #24 OR #25 OR #26 OR #27 OR #28 OR #29 OR #30 OR #31 OR #32 OR #33 OR #34 OR #35 OR #36 OR #37 OR #38 OR #39 OR #40 OR #41 OR #42 OR #43 OR #44 OR #45 OR #46 OR #47 OR #48 OR #49 OR #50 OR #51 OR #52 OR #53 OR #54 OR #55 OR #56 OR #57 OR #58 OR #59 OR #60 OR #61 OR #62 OR #63 OR #64

#64. (polyethylene NEAR/1 glycol):ti,ab

#63. 'heplisav b':ti,ab

#62. ('hepatitis b' NEAR/1 vaccin*):ti,ab

#61. 'hepatitis b vaccine'/exp

#60. dynavax:ti,ab

#59. fluad:ti,ab

#58. pandemrix:ti,ab

#57. arepanrix:ti,ab

#56. nanocrystal*:ti,ab

#55. lnp:ti,ab

#54. nanoparticle*:ti,ab

#53. 'nanoparticle'/exp

#52. saponin*:ti,ab

#51. 'saponin'/exp

#50. (flu NEAR/1 vaccine*):ti,ab

#49. (influenza NEAR/1 vaccine*):ti,ab

#48. 'influenza vaccine'/exp

#47. (vector NEAR/1 expressing):ti,ab

#46. 'mrna lnp':ti,ab

#45. mrna*:ti,ab

#44. bnt162b2:ti,ab

#43. azd1222:ti,ab

#42. v591:ti,ab

#41. 'measles vector':ti,ab

#40. chadox1:ti,ab

#39. 'adenovirus 5':ti,ab

#38. ad5*:ti,ab

#37. 'adenovirus 26':ti,ab

#36. ad26*:ti,ab

#35. (replication NEAR/1 incompetent):ti,ab

#34. (full NEAR/1 length):ti,ab

#33. (molecular NEAR/1 clamp):ti,ab

#32. 's protein*':ti,ab

#31. (stabilized NEAR/1 spike):ti,ab

#30. baculo*:ti,ab

#29. (recombinant NEAR/1 'spike protein'):ti,ab

#28. cpg1:ti,ab

#27. mf59:ti,ab

#26. as03:ti,ab

#25. aluminum*:ti,ab

#24. alhydrogel:ti,ab

#23. 'matrix m*':ti,ab

#22. (immunologic* NEAR/1 adjuvant*):ti,ab

#21. immunoadjuvant*:ti,ab

#20. 'immunological adjuvant'/exp

#19. #1 OR #2 OR #3 OR #4 OR #5 OR #6 OR #7 OR #8 OR #9 OR #10 OR #11 OR #12 OR #13 OR #14 OR #15 OR #16 OR #17 OR #18

#18. dart:ti,ab

#17. fetomaternal:ti,ab

#16. (materno NEAR/1 fetal):ti,ab

#15. maternofetal:ti,ab

#14. fetus:ti,ab

#13. fetal:ti,ab

#12. 'fetus'/exp

#11. partum:ti,ab

#10. parturition*:ti,ab

#9. childbirth*:ti,ab

#8. 'birth'/exp

#7. gestational:ti,ab

#6. miscarriage*:ti,ab

#5. abortion*:ti,ab

#4. 'spontaneous abortion'/exp

#3. 'pregnancy complication'/exp

#2. pregnan*:ti,ab

#1. 'pregnancy'/exp

**Cochrane Library (Wiley)**

#1 MeSH descriptor: [Pregnancy] explode all trees

#2 Pregnan*:ti,ab,kw

#3 MeSH descriptor: [Pregnancy Complications] explode all trees

#4 MeSH descriptor: [Abortion, Spontaneous] explode all trees

#5 Abortion*:ti,ab,kw

#6 Miscarriage*:ti,ab,kw

#7 Gestational:ti,ab,kw

#8 MeSH descriptor: [Parturition] explode all trees

#9 Childbirth*:ti,ab,kw

#10 Parturition*:ti,ab,kw

#11 Partum:ti,ab,kw

#12 MeSH descriptor: [Fetus] explode all trees

#13 Fetal:ti,ab,kw

#14 Fetus:ti,ab,kw

#15 Maternofetal:ti,ab,kw

#16 (Materno NEAR/1 Fetal):ti,ab,kw

#17 Fetomaternal:ti,ab,kw

#18 DART:ti,ab,kw

#19 #1 OR #2 OR #3 OR #4 OR #5 OR #6 OR #7 OR #8 OR #9 OR #10 OR #11 OR #12 OR #13 OR #14 OR #15 OR #16 OR #17 OR #18

#20 MeSH descriptor: [Adjuvants, Immunologic] explode all trees

#21 Immunoadjuvant*:ti,ab,kw

#22 (Immunologic* NEAR/1 Adjuvant*):ti,ab,kw

#23 Matrix-M*:ti,ab,kw

#24 Alhydrogel:ti,ab,kw

#25 Aluminum*:ti,ab,kw

#26 AS03:ti,ab,kw

#27 MF59:ti,ab,kw

#28 CpG1:ti,ab,kw

#29 (Recombinant NEAR/1 Spike-Protein):ti,ab,kw

#30 Baculo*:ti,ab,kw

#31 (Stabilized NEAR/1 Spike):ti,ab,kw

#32 S-Protein*:ti,ab,kw

#33 (Molecular NEAR/1 Clamp):ti,ab,kw

#34 (Full NEAR/1 Length):ti,ab,kw

#35 (Replication NEAR/1 Incompetent):ti,ab,kw

#36 Ad26*:ti,ab,kw

#37 Adenovirus-26:ti,ab,kw

#38 Ad5*:ti,ab,kw

#39 Adenovirus-5:ti,ab,kw

#40 ChAdOx1:ti,ab,kw

#41 (Measles NEAR/1 Vector):ti,ab,kw

#42 V591:ti,ab,kw

#43 AZD1222:ti,ab,kw

#44 BNT162b2:ti,ab,kw

#45 mRNA*:ti,ab,kw

#46 mRNA-LNP:ti,ab,kw

#47 (Vector NEAR/1 Expressing):ti,ab,kw

#48 MeSH descriptor: [Influenza Vaccines] explode all trees

#49 (Influenza NEAR/1 Vaccine*):ti,ab,kw

#50 (Flu NEAR/1 Vaccine*):ti,ab,kw

#51 MeSH descriptor: [Saponins] explode all trees

#52 Saponin*:ti,ab,kw

#53 MeSH descriptor: [Nanoparticles] explode all trees

#54 Nanoparticle*:ti,ab,kw

#55 LNP:ti,ab,kw

#56 Nanocrystal*:ti,ab,kw

#57 Arepanrix:ti,ab,kw

#58 Pandemrix:ti,ab,kw

#59 FLUAD:ti,ab,kw

#60 Dynavax:ti,ab,kw

#61 MeSH descriptor: [Hepatitis B Vaccines] explode all trees

#62 (Hepatitis-B NEAR/1 Vaccin*):ti,ab,kw

#63 HEPLISAV-B:ti,ab,kw

#64 (Polyethylene NEAR/1 Glycol):ti,ab,kw

#65 #20 OR #21 OR #22 OR #23 OR #24 OR #25 OR #26 OR #27 OR #28 OR #29 OR #30 OR #31 OR #32 OR #33 OR #34 OR #35 OR #36 OR #37 OR #38 OR #39 OR #40 OR #41 OR #42 OR #43 OR #44 OR #45 OR #46 OR #47 OR #48 OR #49 OR #50 OR #51 OR #52 OR #53 OR #54 OR #55 OR #56 OR #57 OR #58 OR #59 OR #60 OR #61 OR #62 OR #63 OR #64

#66 MeSH descriptor: [Vaccines] explode all trees

#67 MeSH descriptor: [Vaccination] explode all trees

#68 Vaccin*:ti,ab,kw

#69 #66 OR #67 OR #68

#70 #19 AND #65 AND #69

**CINAHL (EBSCO)**

S85	S72 OR S84

S84	S82 AND S83

S83	(MH "Animals+")

S82	S77 AND S81

S81	S78 OR S79 OR S80

S80	TI DART OR AB DART

S79	TI Pregnan* OR AB Pregnan*

S78	(MH "Pregnancy+")

S77	S73 OR S74 OR S75 OR S76

S76	TI Nanocrystal* OR AB Nanocrystal*

S75	TI LNP OR AB LNP

S74	TI Nanoparticle* OR AB Nanoparticle*

S73	(MH "Nanoparticles")

S72	S19 AND S67 AND S71

S71	S68 OR S69 OR S70

S70	TI Vaccin* OR AB Vaccin*

S69	(MH "Immunization+")

S68	(MH "Vaccines+")

S67	S20 OR S21 OR S22 OR S23 OR S24 OR S25 OR S26 OR S27 OR S28 OR S29 OR S30 OR S31 OR S32 OR S33 OR S34 OR S35 OR S36 OR S37 OR S38 OR S39 OR S40 OR S41 OR S42 OR S43 OR S44 OR S45 OR S46 OR S47 OR S48 OR S49 OR S50 OR S51 OR S52 OR S53 OR S54 OR S55 OR S56 OR S57 OR S58 OR S59 OR S60 OR S61 OR S62 OR S63 OR S64 OR S65 OR S66

S66	TI (Polyethylene N1 Glycol) OR AB (Polyethylene N1 Glycol)

S65	TI HEPLISAV-B OR AB HEPLISAV-B

S64	TI (Hepatitis-B N1 Vaccin*) OR AB (Hepatitis-B N1 Vaccin*)

S63	(MH "Hepatitis B Vaccines+")

S62	TI Dynavax OR AB Dynavax

S61	TI FLUAD OR AB FLUAD

S60	TI Pandemrix OR AB Pandemrix

S59	TI Arepanrix OR AB Arepanrix

S58	TI Nanocrystal* OR AB Nanocrystal*

S57	TI LNP OR AB LNP

S56	TI Nanoparticle* OR AB Nanoparticle*

S55	(MH "Nanoparticles")

S54	TI Saponin* OR AB Saponin*

S53	TI (Flu N1 Vaccine*) OR AB (Flu N1 Vaccine*)

S52	TI (Influenza N1 Vaccine*) OR AB (Influenza N1 Vaccine*)

S51	(MH "Influenza Vaccine")

S50	TI (Vector N1 Expressing) OR AB (Vector N1 Expressing)

S49	TI mRNA-LNP OR AB mRNA-LNP

S48	TI mRNA* OR AB mRNA*

S47	TI BNT162b2 OR AB BNT162b2

S46	TI AZD1222 OR AB AZD1222

S45	TI V591 OR AB V591

S44	TI V591 OR AB V591

S43	TI (Measles N1 Vector) OR AB (Measles N1 Vector)

S42	TI ChAdOx1 OR AB ChAdOx1

S41	TI Adenovirus-5 OR AB Adenovirus-5

S40	TI Ad5* OR AB Ad5*

S39	TI Adenovirus-26 OR AB Adenovirus-26

S38	TI Adenovirus-26 OR AB Adenovirus-26

S37	TI Adenovirus-26 OR AB Adenovirus-26

S36	TI Adenovirus-26 OR AB Adenovirus-26

S35	TI Ad26* OR AB Ad26*

S34	TI (Replication N1 Incompetent) OR AB (Replication N1 Incompetent)

S33	TI (Full N1 Length) OR AB (Full N1 Length)

S32	TI (Molecular N1 Clamp) OR AB (Molecular N1 Clamp)

S31	TI S-Protein* OR AB S-Protein*

S30	TI (Stabilized N1 Spike) OR AB (Stabilized N1 Spike)

S29	TI Baculo* OR AB Baculo*

S28	TI (Recombinant N1 Spike-Protein) OR AB (Recombinant N1 Spike-Protein)

S27	TI CpG1 OR AB CpG1

S26	TI MF59 OR AB MF59

S25	TI AS03 OR AB AS03

S24	TI Aluminum* OR AB Aluminum*

S23	TI Alhydrogel OR AB Alhydrogel

S22	TI Matrix-M* OR AB Matrix-M*

S21	TI (Immunologic* N1 Adjuvant*) OR AB (Immunologic* N1 Adjuvant*)

S20	TI Immunoadjuvant* OR AB Immunoadjuvant*

S19	S1 OR S2 OR S3 OR S4 OR S5 OR S6 OR S7 OR S8 OR S9 OR S10 OR S11 OR S12 OR S13 OR S14 OR S15 OR S16 OR S17 OR S18

S18	TI DART OR AB DART

S17	TI Fetomaternal OR AB Fetomaternal

S16	TI (Materno N1 Fetal) OR AB (Materno N1 Fetal)

S15	TI Maternofetal OR AB Maternofetal

S14	TI Fetus OR AB Fetus

S13	TI Fetal OR AB Fetal

S12	(MH "Fetus+")

S11	TI Partum OR AB Partum

S10	TI Parturition* OR AB Parturition*

S9	TI Childbirth* OR AB Childbirth*

S8	(MH "Labor+")

S7	TI Gestational OR AB Gestational

S6	TI Miscarriage* OR AB Miscarriage*

S5	TI Abortion* OR AB Abortion*

S4	(MH "Abortion, Spontaneous+")

S3	(MH "Pregnancy Complications+")

S2	TI Pregnan* OR AB Pregnan*

S1	(MH "Pregnancy+")

**Global Health (OVID)**

1	exp Pregnancy/

2	Pregnan*.ti,ab.

3	exp Pregnancy Complications/

4	Abortion*.ti,ab.

5	Miscarriage*.ti,ab.

6	Gestational.ti,ab.

7	exp Parturition/

8	Childbirth*.ti,ab.

9	Parturition*.ti,ab.

10	Partum.ti,ab.

11	exp Fetus/

12	Fetal.ti,ab.

13	Fetus.ti,ab.

14	Maternofetal.ti,ab.

15	(Materno adj1 Fetal).ti,ab.

16	Fetomaternal.ti,ab.

17	DART.ti,ab.

18	or/1-17

19	Immunoadjuvant*.ti,ab.

20	(Immunologic* adj1 Adjuvant*).ti,ab.

21	Matrix-M*.mp.

22	Alhydrogel.mp.

23	Aluminum*.mp.

24	AS03.mp.

25	MF59.mp.

26	CpG1.mp.

27	(Recombinant adj1 Spike-Protein).mp.

28	Baculo*.mp.

29	(Stabilized adj1 Spike).mp.

30	S-Protein*.mp.

31	(Molecular adj1 Clamp).mp.

32	(Full adj1 Length).mp.

33	(Replication adj1 Incompetent).mp.

34	Ad26*.mp.

35	Adenovirus-26.mp.

36	Ad5*.mp.

37	Adenovirus-5.mp.

38	ChAdOx1.mp.

39	Measles-Vector.mp.

40	V591.mp.

41	AZD1222.mp.

42	BNT162b2.mp.

43	mRNA*.mp.

44	(Vector adj1 Expressing).mp.

45	(Influenza adj1 Vaccine*).mp.

46	(Flu adj1 Vaccine*).mp.

47	exp Saponins/

48	Saponin*.mp.

49	exp Nanoparticles/

50	Nanoparticle*.mp.

51	LNP.mp.

52	Nanocrystal*.mp.

53	Arepanrix.mp.

54	Pandemrix.mp.

55	FLUAD.mp.

56	Dynavax.mp.

57	(Hepatitis-B adj1 Vaccin*).mp.

58	HEPLISAV-B.mp.

59	(Polyethylene adj1 Glycol).mp.

60	or/19-59

61	exp Vaccination/

62	exp Vaccines/

63	Vaccin*.ti,ab.

64	or/61-63

65	18 and 60 and 64

66	Animal*.mp.

67	exp Nanoparticles/

68	Nanoparticle*.mp.

69	LNP.mp.

70	Nanocrystal*.mp.

71	or/67-70

72	exp Pregnancy/

73	exp Pregnancy/

74	Pregnan*.ti,ab.

75	DART.ti,ab.

76	or/72-74

77	66 and 71 and 76

78	65 or 77

**Appendix 2. Risk of bias assessment tools by study design**

**2.1 Criteria for judging risk of bias in the ‘Risk of bias’ assessment tool**

| **RANDOM SEQUENCE GENERATION**  **Selection bias (biased allocation to interventions) due to inadequate generation of a randomised sequence** | | | |
| --- | --- | --- | --- |
| Criteria for a judgement of ‘Low risk’ of bias. | | The investigators describe a random component in the sequence generation process such as:   - Referring to a random number table; - Using a computer random number generator; - Coin tossing; - Shuffling cards or envelopes; - Throwing dice; - Drawing of lots; - Minimization*.   **Minimization may be implemented without a random element, and this is considered to be equivalent to being random.* | |
| ‘High risk’ of bias. | | The investigators describe a non-random component in the sequence generation process. Usually, the description would involve some systematic, non-random approach, for example:   - Sequence generated by odd or even date of birth; - Sequence generated by some rule based on date (or day) of admission; - Sequence generated by some rule based on hospital or clinic record number.   Other non-random approaches happen much less frequently than the systematic approaches mentioned above and tend to be obvious. They usually involve judgement or some method of non-random categorization of participants, for example:   - Allocation by judgement of the clinician; - Allocation by preference of the participant; - Allocation based on the results of a laboratory test or a series of tests;   Allocation by availability of the intervention. | |
| ‘Unclear risk’ of bias. | | Insufficient information about the sequence generation process to permit judgement of ‘Low risk’ or ‘High risk’. | |
| **ALLOCATION CONCEALMENT**  **Selection bias (biased allocation to interventions) due to inadequate concealment of allocations prior to assignment** | | | |
| ‘Low risk’ of bias. | Participants and investigators enrolling participants could not foresee assignment because one of the following, or an equivalent method, was used to conceal allocation:   - Central allocation (including telephone, web-based and pharmacy-controlled randomization); - Sequentially numbered drug containers of identical appearance; - Sequentially numbered, opaque, sealed envelopes. | | |
| ‘High risk’ of bias. | Participants or investigators enrolling participants could possibly foresee assignments and thus introduce selection bias, such as allocation based on:   - Using an open random allocation schedule (e.g. a list of random numbers); - Assignment envelopes were used without appropriate safeguards (e.g. if envelopes were unsealed or non-opaque or not sequentially numbered); - Alternation or rotation; - Date of birth; - Case record number; - Any other explicitly unconcealed procedure. | | |
| ‘Unclear risk’ of bias. | Insufficient information to permit judgement of ‘Low risk’ or ‘High risk’. This is usually the case if the method of concealment is not described or not described in sufficient detail to allow a definite judgement – for example if the use of assignment envelopes is described, but it remains unclear whether envelopes were sequentially numbered, opaque and sealed. | | |
| **BLINDING OF PARTICIPANTS AND PERSONNEL**  **Performance bias due to knowledge of the allocated interventions by participants and personnel during the study** | | | |
| ‘Low risk’ of bias. | | | Any one of the following:   - No blinding or incomplete blinding, but the review authors judge that the outcome is not likely to be influenced by lack of blinding; - Blinding of participants and key study personnel ensured, and unlikely that the blinding could have been broken. |
| ‘High risk’ of bias. | | | Any one of the following:   - No blinding or incomplete blinding, and the outcome is likely to be influenced by lack of blinding; - Blinding of key study participants and personnel attempted, but likely that the blinding could have been broken, and the outcome is likely to be influenced by lack of blinding. |
| ‘Unclear risk’ of bias. | | | Any one of the following:   - Insufficient information to permit judgement of ‘Low risk’ or ‘High risk’; - The study did not address this outcome. |
| **BLINDING OF OUTCOME ASSESSMENT**  **Detection bias due to knowledge of the allocated interventions by outcome assessors** | | | |
| ‘Low risk’ of bias. | | | Any one of the following:   - No blinding of outcome assessment, but the review authors judge that the outcome measurement is not likely to be influenced by lack of blinding; - Blinding of outcome assessment ensured, and unlikely that the blinding could have been broken. |
| ‘High risk’ of bias. | | | Any one of the following:   - No blinding of outcome assessment, and the outcome measurement is likely to be influenced by lack of blinding; - Blinding of outcome assessment, but likely that the blinding could have been broken, and the outcome measurement is likely to be influenced by lack of blinding. |
| ‘Unclear risk’ of bias. | | | Any one of the following:   - Insufficient information to permit judgement of ‘Low risk’ or ‘High risk’; - The study did not address this outcome. |
| **INCOMPLETE OUTCOME DATA**  **Attrition bias due to amount, nature or handling of incomplete outcome data** | | | |
| ‘Low risk’ of bias. | | | Any one of the following:   - No missing outcome data; - Reasons for missing outcome data unlikely to be related to true outcome (for survival data, censoring unlikely to be introducing bias); - Missing outcome data balanced in numbers across intervention groups, with similar reasons for missing data across groups; - For dichotomous outcome data, the proportion of missing outcomes compared with observed event risk not enough to have a clinically relevant impact on the intervention effect estimate; - For continuous outcome data, plausible effect size (difference in means or standardized difference in means) among missing outcomes not enough to have a clinically relevant impact on observed effect size; - Missing data have been imputed using appropriate methods. |
| ‘High risk’ of bias. | | | Any one of the following:   - Reason for missing outcome data likely to be related to true outcome, with either imbalance in numbers or reasons for missing data across intervention groups; - For dichotomous outcome data, the proportion of missing outcomes compared with observed event risk enough to induce clinically relevant bias in intervention effect estimate; - For continuous outcome data, plausible effect size (difference in means or standardized difference in means) among missing outcomes enough to induce clinically relevant bias in observed effect size; - ‘As-treated’ analysis done with substantial departure of the intervention received from that assigned at randomization; - Potentially inappropriate application of simple imputation. |
| ‘Unclear risk’ of bias. | | | Any one of the following:   - Insufficient reporting of attrition/exclusions to permit judgement of ‘Low risk’ or ‘High risk’ (e.g. number randomized not stated, no reasons for missing data provided); - The study did not address this outcome. |
| **SELECTIVE REPORTING**  **Reporting bias due to selective outcome reporting** | | | |
| ‘Low risk’ of bias. | | | Any of the following:   - The study protocol is available and all of the study’s pre-specified (primary and secondary) outcomes that are of interest in the review have been reported in the pre-specified way; - The study protocol is not available but it is clear that the published reports include all expected outcomes, including those that were pre-specified (convincing text of this nature may be uncommon). |
| ‘High risk’ of bias. | | | Any one of the following:   - Not all of the study’s pre-specified primary outcomes have been reported; - One or more primary outcomes is reported using measurements, analysis methods or subsets of the data (e.g. subscales) that were not pre-specified; - One or more reported primary outcomes were not pre-specified (unless clear justification for their reporting is provided, such as an unexpected adverse effect); - One or more outcomes of interest in the review are reported incompletely so that they cannot be entered in a meta-analysis; - The study report fails to include results for a key outcome that would be expected to have been reported for   such a study. |
| ‘Unclear risk’ of bias. | | | Insufficient information to permit judgement of ‘Low risk’ or ‘High risk’. It is likely that the majority of studies will fall into this category. |
| **OTHER BIAS**  **Bias due to problems not covered elsewhere in the table** | | | |
| ‘Low risk’ of bias. | | | The study appears to be free of other sources of bias. |
| ‘High risk’ of bias. | | | There is at least one important risk of bias. For example, the study:   - Had a potential source of bias related to the specific study design used; or - Has been claimed to have been fraudulent; or - Had some other problem. |
| ‘Unclear’ risk of bias. | | | There may be a risk of bias, but there is either:   - Insufficient information to assess whether an important risk of bias exists; or - Insufficient rationale or evidence that an identified problem will introduce bias. |

**2.2 Criteria for judging risk of bias in quasi-experimental studies (‘Cochrane EPOC’ assessment tool**

**QUALITY CRITERIA FOR CONTROLLED BEFORE AND AFTER (CBA) DESIGNS**

Seven standard criteria are used for CBAs included in EPOC reviews:

**a) Baseline measurement:**

LOW RISK if performance or patient outcomes were measured prior to the intervention, and no substantial differences were present across study groups (e.g. where multiple pre-intervention measures describe similar trends in intervention and control groups);

UNCLEAR RISK if baseline measures are not reported, or if it is unclear whether baseline measures are substantially different across study groups;

HIGH RISK if there are differences at baseline in main outcome measures likely to undermine the post-intervention differences (e.g. are differences between the groups before the intervention similar to those found post-intervention).

**b) Characteristics for studies using second site as control:**

LOW RISK if characteristics of study and control providers are reported and similar;

UNCLEAR RISK if it is not clear in the paper e.g. characteristics are mentioned in the text but no data are presented;

HIGH RISK if there is no report of characteristics either in the text or a table OR if baseline characteristics are reported and there are differences between study and control providers.

**c) Blinded assessment of primary outcome(s)* (protection against detection bias):**

LOW RISK if the authors state explicitly that the primary outcome variables were assessed blindly OR the outcome variables are objective e.g. length of hospital stay, drug levels as assessed by a standardised test;

UNCLEAR RISK if not specified in the paper;

HIGH RISK if the outcomes were not assessed blindly.

** Primary outcome(s) are those variables that correspond to the primary hypothesis or question as defined by the authors. In the event that some of the primary outcome variables were assessed in a blind fashion and others were not, score each separately and label each outcome variable clearly.*

**d) Protection against contamination:**

*Studies using second site as control:*

LOW RISK if allocation was by community, institution, or practice and is unlikely that the control group received the intervention;

UNCLEAR RISK if providers were allocated within a clinic or practice and communication between experimental and group providers was likely to occur;

HIGH RISK if it is likely that the control group received the intervention (e.g. cross-over studies or if patients rather than providers were randomised).

**e) Reliable primary outcome measure(s):**

LOW RISK if two or more raters with at least 90% agreement or kappa greater than or equal to 0.8 OR the outcome is obtained from some automated system e.g. length of hospital stay, drug levels as assessed by a standardised test;

UNCLEAR RISK if reliability is not reported for outcome measures that are obtained by chart extraction or collected by an individual;

HIGH RISK if agreement is less than 90% or kappa is less than 0.8.

** In the event that some outcome variables were assessed in a reliable fashion and others were not, score each separately and label each outcome variable clearly.*

**f) Follow-up of professionals (protection against exclusion bias):**

LOW RISK if outcome measures obtained 80-100% subjects allocated to groups. (Do not assume 100% follow-up unless stated explicitly.);

UNCLEAR RISK if not specified in the paper;

HIGH RISK if outcome measures obtained for less than 80% of patients allocated to groups.

**g) Follow-up of patients:**

LOW RISK if outcome measures obtained 80-100% of patients allocated to groups or for patients who entered the study. (Do not assume 100% follow-up unless stated explicitly.);

UNCLEAR RISK if not specified in the paper;

HIGH RISK if outcome measures obtained for less than 80% of patients allocated to groups or for less than 80% of patients who entered the study.

**QUALITY CRITERIA FOR INTERRUPTED TIME SERIES (ITS)**

The following seven standard criteria should be used to assess the methodology quality of ITS designs included in EPOC reviews. Each criterion is scored DONE, NOT CLEAR or NOT DONE but here we use 'low risk', 'unclear risk', and 'high risk' respectively to be consistent with the 'Risk of bias' assessment tool for RCTs (Appendix 2.1).

*Protection against secular changes:*

**a) The intervention is independent of other changes.**

LOW RISK if the intervention occurred independently of other changes over time;

UNCLEAR RISK if not specified (will be treated as HIGH RISK if information cannot be obtained from the authors);

HIGH RISK if reported that intervention was not independent of other changes in time.

**b) Data were analysed appropriately:**

LOW RISK if ARIMA models were used OR time series regression models were used to analyse the data and serial correlation was adjusted or tested for;

UNCLEAR RISK if not specified (will be treated as HIGH RISK if information cannot be obtained from the authors);

HIGH RISK if it is clear that neither of the conditions above not met.

**c) Reason for the number of points pre- and post-intervention given:**

LOW RISK if rationale for the number of points stated (e.g. monthly data for 12 months post-intervention was used because the anticipated effect was expected to decay) OR sample size calculation performed;

UNCLEAR RISK if not specified (will be treated as HIGH RISK if information cannot be obtained from the authors);

HIGH RISK if it is clear that neither of the conditions above met.

**d) Shape of the intervention effect was specified:**

LOW RISK if a rational explanation for the shape of intervention effect was given by the author(s);

UNCLEAR RISK if not specified (will be treated as HIGH RISK if information cannot be obtained from the authors);

HIGH RISK if it is clear that the condition above is not met.

*Protection against detection bias:*

**e) Intervention unlikely to affect data collection:**

LOW RISK if reported that intervention itself was unlikely to affect data collection (for example, sources and methods of data collection were the same before and after the intervention);

UNCLEAR RISK if not reported (will be treated as HIGH RISK if information cannot be obtained from the authors);

HIGH RISK if the intervention itself was likely to affect data collection (for example, any change in source or method of data collection reported).

**f) Blinded assessment of primary outcome(s)*:**

LOW RISK if the authors state explicitly that the primary outcome variables were assessed blindly OR the outcome variables are objective e.g. length of hospital stay, drug levels as assessed by a standardised test;

UNCLEAR RISK if not specified (will be treated as HIGH RISK if information cannot be obtained from the authors);

HIGH RISK if the outcomes were not assessed blindly.

** Primary outcome(s) are those variables that correspond to the primary hypothesis or question as defined by the authors. In the event that some of the primary outcome variables were assessed in a blind fashion and others were not, score each separately and label each outcome variable clearly.*

**g) Completeness of data set:**

LOW RISK if data set covers 80-100% of total number of participants or episodes of care in the study;

UNCLEAR RISK if not specified (will be treated as HIGH RISK if information cannot be obtained from the authors);

HIGH RISK if data set covers less than 80% of the total number of participants or episodes of care in the study.

**h) Reliable primary outcome measure(s)*:**

LOW RISK if two or more raters with at least 90% agreement or kappa greater than or equal to 0.8 OR the outcome is obtained from some automated system e.g. length of hospital stay, drug levels as assessed by a standardised test;

UNCLEAR RISK if reliability is not reported for outcome measures that are obtained by chart extraction or collected by an individual (will be treated as HIGH RISK if information cannot be obtained from the authors);

HIGH RISK if agreement is less than 90% or kappa is less than 0.8.

** In the event that some outcome variables were assessed in a reliable fashion and others were not, score each separately.*

**QUALITY CRITERIA FOR CONTROLLED INTERRUPTED TIME SERIES (CITS)**

**a) Protection against secular changes:**

The intervention is independent of other changes.

LOW RISK if the intervention occurred independently of other changes over time;

UNCLEAR RISK if not specified (will be treated as HIGH RISK if information cannot be obtained from the authors);

HIGH RISK if reported that intervention was not independent of other changes in time.

**b) Data were analysed appropriately:**

LOW RISK if ARIMA models were used OR time series regression models were used to analyse the data and serial correlation was adjusted or tested for;

UNCLEAR RISK if not specified (will be treated as HIGH RISK if information cannot be obtained from the authors);

HIGH RISK if it is clear that neither of the conditions above not met.

**c) Reason for the number of points pre- and post-intervention given:**

LOW RISK if rationale for the number of points stated (e.g. monthly data for 12 months post-intervention was used because the anticipated effect was expected to decay) OR sample size calculation performed;

UNCLEAR RISK if not specified (will be treated as HIGH RISK if information cannot be obtained from the authors);

HIGH RISK if it is clear that neither of the conditions above met.

**d) Shape of the intervention effect was specified:**

LOW RISK if a rational explanation for the shape of intervention effect was given by the author(s);

UNCLEAR RISK if not specified (will be treated as HIGH RISK if information cannot be obtained from the authors);

HIGH RISK if it is clear that the condition above is not met.

*Intervention unlikely to affect data collection:*

**e) Protection against detection bias:**

LOW RISK if reported that intervention itself was unlikely to affect data collection (for example, sources and methods of data collection were the same before and after the intervention);

UNCLEAR RISK if not reported (will be treated as HIGH RISK if information cannot be obtained from the authors);

HIGH RISK if the intervention itself was likely to affect data collection (for example, any change in source or method of data collection reported).

**f) Blinded assessment of primary outcome(s)*:**

LOW RISK if the authors state explicitly that the primary outcome variables were assessed blindly OR the outcome variables are objective e.g. length of hospital stay, drug levels as assessed by a standardised test;

UNCLEAR RISK if not specified (will be treated as HIGH RISK if information cannot be obtained from the authors);

HIGH RISK if the outcomes were not assessed blindly.

* Primary outcome(s) are those variables that correspond to the primary hypothesis or question as defined by the authors. In the event that some of the primary outcome variables were assessed in a blind fashion and others were not, score each separately and label each outcome variable clearly.

**g) Completeness of data set:**

LOW RISK if data set covers 80-100% of total number of participants or episodes of care in the study;

UNCLEAR RISK if not specified (will be treated as HIGH RISK if information cannot be obtained from the authors);

HIGH RISK if data set covers less than 80% of the total number of participants or episodes of care in the study.

**h) Reliable primary outcome measure(s)*:**

LOW RISK if two or more raters with at least 90% agreement or kappa greater than or equal to 0.8 OR the outcome is obtained from some automated system e.g. length of hospital stay, drug levels as assessed by a standardised test;

UNCLEAR RISK if reliability is not reported for outcome measures that are obtained by chart extraction or collected by an individual (will be treated as HIGH RISK if information cannot be obtained from the authors);

HIGH RISK if agreement is less than 90% or kappa is less than 0.8.

* In the event that some outcome variables were assessed in a reliable fashion and others were not, score each separately.

For CITSs, as for CBAs, we will include three additional domains that assess design-specific threats to validity covered by the Cochrane EPOC group: imbalance of outcome measures at baseline; comparability of intervention and control group characteristics at baseline; and protection against contamination.

**i) Baseline measurement:**

LOW RISK if performance or patient outcomes were measured prior to the intervention, and no substantial differences were present across study groups (e.g. where multiple pre-intervention measures describe similar trends in intervention and control groups);

UNCLEAR RISK if baseline measures are not reported, or if it is unclear whether baseline measures are substantially different across study groups;

HIGH RISK if there are differences at baseline in main outcome measures likely to undermine the post-intervention differences (e.g. are differences between the groups before the intervention similar to those found post-intervention).

**j) Characteristics for studies using second site as control:**

LOW RISK if characteristics of study and control providers are reported and similar;

UNCLEAR RISK if it is not clear in the paper e.g. characteristics are mentioned in the text but no data are presented;

HIGH RISK if there is no report of characteristics either in the text or a table OR if baseline characteristics are reported and there are differences between study and control providers.

*Studies using second site as control:*

**k) Protection against contamination:**

LOW RISK if allocation was by community, institution, or practice and is unlikely that the control group received the intervention;

UNCLEAR RISK if providers were allocated within a clinic or practice and communication between experimental and group providers was likely to occur;

HIGH RISK if it is likely that the control group received the intervention (e.g. cross-over studies or if patients rather than providers were randomised).

**QUALITY CRITERIA FOR UNCONTROLLED BEFORE AND AFTER (UBA) DESIGNS**

Four standard criteria are used for UBAs (Derived from CBAs EPOC criteria):

**a) Blinded assessment of primary outcome(s)* (protection against detection bias):**

LOW RISK if the authors state explicitly that the primary outcome variables were assessed blindly OR the outcome variables are objective e.g. length of hospital stay, drug levels as assessed by a standardised test;

UNCLEAR RISK if not specified in the paper;

HIGH RISK if the outcomes were not assessed blindly.

** Primary outcome(s) are those variables that correspond to the primary hypothesis or question as defined by the authors. In the event that some of the primary outcome variables were assessed in a blind fashion and others were not, score each separately and label each outcome variable clearly.*

**b) Reliable primary outcome measure(s):**

LOW RISK if two or more raters with at least 90% agreement or kappa greater than or equal to 0.8 OR the outcome is obtained from some automated system e.g. length of hospital stay, drug levels as assessed by a standardised test;

UNCLEAR RISK if reliability is not reported for outcome measures that are obtained by chart extraction or collected by an individual;

HIGH RISK if agreement is less than 90% or kappa is less than 0.8.

** In the event that some outcome variables were assessed in a reliable fashion and others were not, score each separately and label each outcome variable clearly.*

**c) Follow-up of professionals (protection against exclusion bias):**

LOW RISK if outcome measures obtained 80-100% subjects at baseline. (Do not assume 100% follow-up unless stated explicitly.);

UNCLEAR RISK if not specified in the paper;

HIGH RISK if outcome measures obtained for less than 80% of patients at baseline.

**d) Follow-up of patients:**

LOW RISK if outcome measures obtained 80-100% of patients who entered the study. (Do not assume 100% follow-up unless stated explicitly.);

UNCLEAR RISK if not specified in the paper;

HIGH RISK if outcome measures obtained for less than 80% of patients who entered the study.

**2.3 NIH Quality Assessment Tool for observational studies**

<https://www.nhlbi.nih.gov/health-topics/study-quality-assessment-tools>

| **Criteria for cohort and cross-sectional studies** | **Judgement*** |
| --- | --- |
| 1. Was the research question or objective in this paper clearly stated? |  |
| 2. Was the study population clearly specified and defined? |  |
| 3. Was the participation rate of eligible persons at least 50%? |  |
| 4. Were all the subjects selected or recruited from the same or similar populations (including the same time period)? Were inclusion and exclusion criteria for being in the study prespecified and applied uniformly to all participants? |  |
| 5. Was a sample size justification, power description, or variance and effect estimates provided? |  |
| 6. For the analyses in this paper, were the exposure(s) of interest measured prior to the outcome(s) being measured? |  |
| 7. Was the timeframe sufficient so that one could reasonably expect to see an association between exposure and outcome if it existed? |  |
| 8. For exposures that can vary in amount or level, did the study examine different levels of the exposure as related to the outcome (e.g., categories of exposure, or exposure measured as continuous variable)? |  |
| 9. Were the exposure measures (independent variables) clearly defined, valid, reliable, and implemented consistently across all study participants? |  |
| 10. Was the exposure(s) assessed more than once over time? |  |
| 11. Were the outcome measures (dependent variables) clearly defined, valid, reliable, and implemented consistently across all study participants? |  |
| 12. Were the outcome assessors blinded to the exposure status of participants? |  |
| 13. Was loss to follow-up after baseline 20% or less? |  |
| 14. Were key potential confounding variables measured and adjusted statistically for their impact on the relationship between exposure(s) and outcome(s)? |  |

***Yes, No, CD, cannot determine; NA, not applicable; NR, not reported**

| **Criteria for cohort and case-control studies** | **Judgement*** |
| --- | --- |
| 1. Was the research question or objective in this paper clearly stated and appropriate? |  |
| 2. Was the study population clearly specified and defined? |  |
| 3. Did the authors include a sample size justification? |  |
| 4. Were controls selected or recruited from the same or similar population that gave rise to the cases (including the same timeframe)? |  |
| 5. Were the definitions, inclusion and exclusion criteria, algorithms or processes used to identify or select cases and controls valid, reliable, and implemented consistently across all study participants? |  |
| 6. Were the cases clearly defined and differentiated from controls? |  |
| 7. If less than 100 percent of eligible cases and/or controls were selected for the study, were the cases and/or controls randomly selected from those eligible? |  |
| 8. Was there use of concurrent controls? |  |
| 9. Were the investigators able to confirm that the exposure/risk occurred prior to the development of the condition or event that defined a participant as a case? |  |
| 10. Were the measures of exposure/risk clearly defined, valid, reliable, and implemented consistently (including the same time period) across all study participants? |  |
| 11. Were the assessors of exposure/risk blinded to the case or control status of participants? |  |
| 12. Were key potential confounding variables measured and adjusted statistically in the analyses? If matching was used, did the investigators account for matching during study analysis? |  |

***Yes, No, CD, cannot determine; NA, not applicable; NR, not reported**

| **Criteria for case-series studies** | **Judgement*** |
| --- | --- |
| 1. Was the study question or objective clearly stated? |  |
| 2. Was the study population clearly and fully described, including a case definition? |  |
| 3. Were the cases consecutive? |  |
| 4. Were the subjects comparable? |  |
| 5. Was the intervention clearly described? |  |
| 6. Were the outcome measures clearly defined, valid, reliable, and implemented consistently across all study participants? |  |
| 7. Was the length of follow-up adequate? |  |
| 8. Were the statistical methods well-described? |  |
| 9. Were the results well-described? |  |

***Yes, No, CD, cannot determine; NA, not applicable; NR, not reported**

**Appendix 3. List and exclusion reasons of excluded studies**

| Author | Title | Exclusion reason |
| --- | --- | --- |
| Alguacil-Ramos 2015^1^ | [Safety of influenza vaccines in risk groups: analysis of adverse events following immunization reported in Valencian Community from 2005 to 2011] | Wrong intervention |
| Arriola 2017^2^ | Association of influenza vaccination during pregnancy with birth outcomes in Nicaragua | Wrong intervention |
| Beau 2014^3^ | Pandemic A/H1N1 influenza vaccination during pregnancy: a comparative study using the EFEMERIS database | Wrong intervention |
| Carcione 2013^4^ | Safety surveillance of influenza vaccine in pregnant women | Wrong intervention |
| Chambers 2013^5^ | Risks and safety of pandemic H1N1 influenza vaccine in pregnancy: birth defects, spontaneous abortion, preterm delivery, and small for gestational age infants | Wrong intervention |
| Chambers 2015^6^ | Safety of seasonal influenza vaccines in pregnancy: VAMPSS update | Wrong intervention |
| Chambers 2016^7^ | Safety of the 2010-11, 2011-12, 2012-13, and 2013-14 seasonal influenza vaccines in pregnancy: birth defects, spontaneous abortion, preterm delivery, and small for gestational age infants, a study from the cohort arm of VAMPSS | Wrong intervention |
| Choe 2011^8^ | Active surveillance of adverse events following immunization against pandemic influenza A (H1N1) in Korea | Wrong intervention |
| Conlin 2013^9^ | Safety of the pandemic H1N1 influenza vaccine among pregnant U.S. military women and their newborns | Wrong intervention |
| Covington 2019^10^ | PIN66 VACCINE PREGNANCY REGISTRIES: METHODS AND IMPACT ON FINDINGS | Wrong intervention |
| Cross 2020^11^ | Adverse events of interest vary by influenza vaccine type and brand: Sentinel network study of eight seasons (2010-2018) | Wrong patient population |
| Dodds 2011^12^ | Influenza vaccination in pregnancy | Wrong intervention |
| Donahue 2017^13^ | Association of spontaneous abortion with receipt of inactivated influenza vaccine containing H1N1pdm09 in 2010-11 and 2011-12 | Wrong intervention |
| Donahue 2019^14^ | Inactivated influenza vaccine and spontaneous abortion in the Vaccine Safety Datalink in 2012-13, 2013-14, and 2014-15 | Wrong intervention |
| Durrieu 2011^15^ | Safety surveillance of influenza A(H1N1)v monovalent vaccines during the 2009-2010 mass vaccination campaign in France | Wrong intervention |
| Eaton 2018^16^ | Birth outcomes following immunization of pregnant women with pandemic H1N1 influenza vaccine 2009-2010 | Wrong intervention |
| Goldman 2013^17^ | Comparison of VAERS fetal-loss reports during three consecutive influenza seasons: was there a synergistic fetal toxicity associated with the two-vaccine 2009/2010 season? | Wrong intervention |
| Kankawinpong 2012^18^ | Immunogenicity and safety of an inactivated pandemic H1N1 vaccine provided by the Thai ministry of public health as a routine public health service | Wrong intervention |
| Kharbanda 2013^19^ | Inactivated influenza vaccine during pregnancy and risks for adverse obstetric events | Wrong intervention |
| Kharbanda 2017^20^ | First trimester influenza vaccination and risks for major structural birth defects in offspring | Wrong intervention |
| Kozuki 2018^21^ | Impact of maternal vaccination timing and influenza virus circulation on birth outcomes in rural Nepal | Wrong intervention |
| Louik 2013^22^ | Risks and safety of pandemic H1N1 influenza vaccine in pregnancy: exposure prevalence, preterm delivery, and specific birth defects | Wrong intervention |
| Louik 2016^23^ | Safety of the 2011-12, 2012-13, and 2013-14 seasonal influenza vaccines in pregnancy: preterm delivery and specific malformations, a study from the case-control arm of VAMPSS | Wrong intervention |
| Lylianou 2012^24^ | Adverse events following immunization from pandemic influenza A (H1N1)-Laos 2010 | Wrong intervention |
| McHugh 2017^25^ | Birth outcomes for Australian mother-infant pairs who received an influenza vaccine during pregnancy, 2012-2014: the FluMum study | Wrong intervention |
| McHugh 2019^26^ | Influenza vaccination in pregnancy among a group of remote dwelling Aboriginal and Torres Strait Islander mothers in the Northern Territory: The 1+1 Healthy Start to Life study | Wrong intervention |
| Mohammed 2020^27^ | Safety and protective effects of maternal influenza vaccination on pregnancy and birth outcomes: A prospective cohort study | Wrong intervention |
| Moro 2017^28^ | Surveillance of Adverse Events After Seasonal Influenza Vaccination in Pregnant Women and Their Infants in the Vaccine Adverse Event Reporting System, July 2010-May 2016 | Wrong intervention |
| Moro 2020^29^ | Monitoring the safety of high-dose, trivalent inactivated influenza vaccine in the vaccine adverse event reporting system (VAERS), 2011 - 2019 | Wrong intervention |
| Nordin 2013^30^ | Maternal safety of trivalent inactivated influenza vaccine in pregnant women | Wrong intervention |
| Ohfuji 2020^31^ | Safety of influenza vaccination on adverse birth outcomes among pregnant women: A prospective cohort study in Japan | Wrong intervention |
| Omer 2011^32^ | Maternal influenza immunization and reduced likelihood of prematurity and small for gestational age births: a retrospective cohort study | Wrong intervention |
| Peppa 2020^33^ | Seasonal influenza vaccination during pregnancy and the risk of major congenital malformations in live-born infants: A 2010-2016 historical cohort study | Wrong intervention |
| Phengxay 2015^34^ | Introducing seasonal influenza vaccine in low-income countries: an adverse events following immunization survey in the Lao People's Democratic Republic | Wrong intervention |
| Regan 2014^35^ | Using SMS to monitor adverse events following trivalent influenza vaccination in pregnant women | Wrong intervention |
| Regan 2016^36^ | Seasonal trivalent influenza vaccination during pregnancy and the incidence of stillbirth: population-based retrospective cohort study | Wrong intervention |
| Regan 2018^37^ | Birth outcomes associated with seasonal influenza vaccination during first trimester of pregnancy | Wrong intervention |
| Richner 2017^38^ | Vaccine mediated protection against Zika virus-induced congenital disease | Wrong outcomes |
| Shatla 2016^39^ | Effect of maternal antenatal influenza vaccination on adverse neonatal outcomes in terms of premature birth, small-for-gestational age and low birth weight: a comparative study | Wrong intervention |
| Vazquez-Benitez 2016^40^ | Risk of preterm or small-for-gestational-age birth after influenza vaccination during pregnancy: caveats when conducting retrospective observational studies | Wrong intervention |
| Walsh 2019^41^ | Health outcomes of young children born to mothers who received 2009 pandemic H1N1 influenza vaccination during pregnancy: retrospective cohort study | Wrong intervention |
| Wijnans 2017^42^ | Bell's palsy and influenza(H1N1)pdm09 containing vaccines: a self-controlled case series | Wrong patient population |

**Appendix 4. PRISMA checklist**

| **Section and Topic** | **Item #** | **Checklist item** | **Location where item is reported** |
| --- | --- | --- | --- |
| **TITLE** | | |  |
| Title | 1 | Identify the report as a systematic review. | 1 |
| **ABSTRACT** | | |  |
| Abstract | 2 | See the PRISMA 2020 for Abstracts checklist. | 1-2 |
| **INTRODUCTION** | | |  |
| Rationale | 3 | Describe the rationale for the review in the context of existing knowledge. | 2-3 |
| Objectives | 4 | Provide an explicit statement of the objective(s) or question(s) the review addresses. | 4 |
| **METHODS** | | |  |
| Eligibility criteria | 5 | Specify the inclusion and exclusion criteria for the review and how studies were grouped for the syntheses. | 4-5 |
| Information sources | 6 | Specify all databases, registers, websites, organisations, reference lists and other sources searched or consulted to identify studies. Specify the date when each source was last searched or consulted. | 5 |
| Search strategy | 7 | Present the full search strategies for all databases, registers and websites, including any filters and limits used. | 5 and Appendix 1 |
| Selection process | 8 | Specify the methods used to decide whether a study met the inclusion criteria of the review, including how many reviewers screened each record and each report retrieved, whether they worked independently, and if applicable, details of automation tools used in the process. | 6 |
| Data collection process | 9 | Specify the methods used to collect data from reports, including how many reviewers collected data from each report, whether they worked independently, any processes for obtaining or confirming data from study investigators, and if applicable, details of automation tools used in the process. | 6 |
| Data items | 10a | List and define all outcomes for which data were sought. Specify whether all results that were compatible with each outcome domain in each study were sought (e.g. for all measures, time points, analyses), and if not, the methods used to decide which results to collect. | 6 |
|  | 10b | List and define all other variables for which data were sought (e.g. participant and intervention characteristics, funding sources). Describe any assumptions made about any missing or unclear information. | 6 |
| Study risk of bias assessment | 11 | Specify the methods used to assess risk of bias in the included studies, including details of the tool(s) used, how many reviewers assessed each study and whether they worked independently, and if applicable, details of automation tools used in the process. | Appendix 2 |
| Effect measures | 12 | Specify for each outcome the effect measure(s) (e.g. risk ratio, mean difference) used in the synthesis or presentation of results. | 6 |
| Synthesis methods | 13a | Describe the processes used to decide which studies were eligible for each synthesis (e.g. tabulating the study intervention characteristics and comparing against the planned groups for each synthesis (item #5)). | 6-7 |
|  | 13b | Describe any methods required to prepare the data for presentation or synthesis, such as handling of missing summary statistics, or data conversions. | 6 |
|  | 13c | Describe any methods used to tabulate or visually display results of individual studies and syntheses. | 6 |
|  | 13d | Describe any methods used to synthesize results and provide a rationale for the choice(s). If meta-analysis was performed, describe the model(s), method(s) to identify the presence and extent of statistical heterogeneity, and software package(s) used. | NA |
|  | 13e | Describe any methods used to explore possible causes of heterogeneity among study results (e.g. subgroup analysis, meta-regression). | NA |
|  | 13f | Describe any sensitivity analyses conducted to assess robustness of the synthesized results. | NA |
| Reporting bias assessment | 14 | Describe any methods used to assess risk of bias due to missing results in a synthesis (arising from reporting biases). | Not described |
| Certainty assessment | 15 | Describe any methods used to assess certainty (or confidence) in the body of evidence for an outcome. | Not described |
| **RESULTS** | | |  |
| Study selection | 16a | Describe the results of the search and selection process, from the number of records identified in the search to the number of studies included in the review, ideally using a flow diagram. | Figure 1 |
|  | 16b | Cite studies that might appear to meet the inclusion criteria, but which were excluded, and explain why they were excluded. | Appendix 3 |
| Study characteristics | 17 | Cite each included study and present its characteristics. | Table 2, 3 and 4 |
| Risk of bias in studies | 18 | Present assessments of risk of bias for each included study. | Table 5 and 6 |
| Results of individual studies | 19 | For all outcomes, present, for each study: (a) summary statistics for each group (where appropriate) and (b) an effect estimate and its precision (e.g. confidence/credible interval), ideally using structured tables or plots. | Table 2 and 3 |
| Results of syntheses | 20a | For each synthesis, briefly summarise the characteristics and risk of bias among contributing studies. | NA |
|  | 20b | Present results of all statistical syntheses conducted. If meta-analysis was done, present for each the summary estimate and its precision (e.g. confidence/credible interval) and measures of statistical heterogeneity. If comparing groups, describe the direction of the effect. | NA |
|  | 20c | Present results of all investigations of possible causes of heterogeneity among study results. | Not described |
|  | 20d | Present results of all sensitivity analyses conducted to assess the robustness of the synthesized results. | Not described |
| Reporting biases | 21 | Present assessments of risk of bias due to missing results (arising from reporting biases) for each synthesis assessed. | NA |
| Certainty of evidence | 22 | Present assessments of certainty (or confidence) in the body of evidence for each outcome assessed. | Not described |
| **DISCUSSION** | | |  |
| Discussion | 23a | Provide a general interpretation of the results in the context of other evidence. | 17 |
|  | 23b | Discuss any limitations of the evidence included in the review. | 18 |
|  | 23c | Discuss any limitations of the review processes used. | 18 |
|  | 23d | Discuss implications of the results for practice, policy, and future research. | 18-19 |
| **OTHER INFORMATION** | | |  |
| Registration and protocol | 24a | Provide registration information for the review, including register name and registration number, or state that the review was not registered. | 7 |
|  | 24b | Indicate where the review protocol can be accessed, or state that a protocol was not prepared. | 7 |
|  | 24c | Describe and explain any amendments to information provided at registration or in the protocol. | NA |
| Support | 25 | Describe sources of financial or non-financial support for the review, and the role of the funders or sponsors in the review. | 19 |
| Competing interests | 26 | Declare any competing interests of review authors. | 19 |
| Availability of data, code and other materials | 27 | Report which of the following are publicly available and where they can be found: template data collection forms; data extracted from included studies; data used for all analyses; analytic code; any other materials used in the review. | NA |

**References**

1.	Alguacil-Ramos AM, Garrigues-Pelufo TM, Muelas-Tirado J, Portero-Alonso A, Perez-Panades J, Fons-Martinez J. [Safety of influenza vaccines in risk groups: analysis of adverse events following immunization reported in Valencian Community from 2005 to 2011]. *Seguridad de las vacunas antigripales en grupos de riesgo: analisis de las sospechas de reacciones adversas notificadas en Comunidad Valenciana entre 2005 y 2011.* 2015;28(4):193-199.

2.	Arriola CS, Vasconez N, Thompson MG, et al. Association of influenza vaccination during pregnancy with birth outcomes in Nicaragua. *Vaccine.* 2017;35(23):3056-3063.

3.	Beau AB, Hurault-Delarue C, Vidal S, et al. Pandemic A/H1N1 influenza vaccination during pregnancy: a comparative study using the EFEMERIS database. *Vaccine.* 2014;32(11):1254-1258.

4.	Carcione D, Blyth CC, Richmond PC, Mak DB, Effler PV. Safety surveillance of influenza vaccine in pregnant women. *The Australian & New Zealand journal of obstetrics & gynaecology.* 2013;53(1):98-99.

5.	Chambers CD, Johnson D, Xu RH, et al. Risks and safety of pandemic H1N1 influenza vaccine in pregnancy: birth defects, spontaneous abortion, preterm delivery, and small for gestational age infants. *Vaccine.* 2013;31(44):5026-5032.

6.	Chambers CD, Louik C, Jones KL, Mitchell AA, Schatz M. Safety of seasonal influenza vaccines in pregnancy: VAMPSS update. *Pharmacoepidemiology and Drug Safety.* 2015;24:12.

7.	Chambers CD, Johnson DL, Xu R, et al. Safety of the 2010-11, 2011-12, 2012-13, and 2013-14 seasonal influenza vaccines in pregnancy: birth defects, spontaneous abortion, preterm delivery, and small for gestational age infants, a study from the cohort arm of VAMPSS. *Vaccine.* 2016;34(37):4443-4449.

8.	Choe YJ, Cho H, Song KM, et al. Active surveillance of adverse events following immunization against pandemic influenza A (H1N1) in Korea. *Japanese journal of infectious diseases.* 2011;64(4):297-303.

9.	Conlin AMS, Bukowinski AT, Sevick CJ, DeScisciolo C, Crum-Cianflone NF. Safety of the pandemic H1N1 influenza vaccine among pregnant U.S. military women and their newborns. *Obstetrics & Gynecology (New York).* 2013;121(3):511-518.

10.	Covington D, Kaydo S, Veley K. Hepatitis B virus (HBV) vaccine in pregnancy and impact on pregnancy outcome. *Value in Health.* 2018;21:S151.

11.	Cross JW, Joy M, McGee C, Akinyemi O, Gatenby P, Lusignan Sd. Adverse events of interest vary by influenza vaccine type and brand: Sentinel network study of eight seasons (2010-2018). *Vaccine.* 2020;38(22):3869-3880.

12.	Dodds L, McNeil S, Scott J, Allen VM, Spencer A, MacDonald N. Influenza vaccination in pregnancy. *American Journal of Epidemiology.* 2011;173:S40.

13.	Donahue JG, Kieke BA, King JP, et al. Association of spontaneous abortion with receipt of inactivated influenza vaccine containing H1N1pdm09 in 2010-11 and 2011-12. *Vaccine.* 2017;35(40):5314-5322.

14.	Donahue JG, Kieke BA, King JP, et al. Inactivated influenza vaccine and spontaneous abortion in the Vaccine Safety Datalink in 2012-13, 2013-14, and 2014-15. *Vaccine.* 2019;37(44):6673-6681.

15.	Durrieu G, Caillet C, Faucher A, et al. Safety surveillance of influenza A(H1N1)v monovalent vaccines during the 2009-2010 mass vaccination campaign in France. *Fundamental and Clinical Pharmacology.* 2011;25:72-73.

16.	Eaton A, Lewis N, Fireman B, et al. Birth outcomes following immunization of pregnant women with pandemic H1N1 influenza vaccine 2009-2010. *Vaccine.* 2018;36(19):2733-2739.

17.	Goldman GS. Comparison of VAERS fetal-loss reports during three consecutive influenza seasons: was there a synergistic fetal toxicity associated with the two-vaccine 2009/2010 season? *Human & Experimental Toxicology.* 2013;32(5):464-475.

18.	Kankawinpong O, Sangsajja C, Cholapand A, et al. Immunogenicity and safety of an inactivate pandemic H1N1 vaccine provided by the Thai ministry of public health as a routine public health service. *The Southeast Asian journal of tropical medicine and public health.* 2012;43(3):680-686.

19.	Kharbanda EO, Vazquez-Benitez G, Lipkind H, Naleway A, Lee G, Nordin JD. Inactivated influenza vaccine during pregnancy and risks for adverse obstetric events. *Obstetrics & Gynecology (New York).* 2013;122(3):659-667.

20.	Kharbanda EO, Vazquez-Benitez G, Romitti PA, et al. First trimester influenza vaccination and risks for major structural birth defects in offspring. *Journal of Pediatrics.* 2017;187:234-e234.

21.	Kozuki N, Katz J, Englund JA, et al. Impact of maternal vaccination timing and influenza virus circulation on birth outcomes in rural Nepal. *International Journal of Gynecology & Obstetrics.* 2018;140(1):65-72.

22.	Louik C, Ahrens K, Kerr S, et al. Risks and safety of pandemic H1N1 influenza vaccine in pregnancy: exposure prevalence, preterm delivery, and specific birth defects. *Vaccine.* 2013;31(44):5033-5040.

23.	Louik C, Kerr S, Bennekom CMv, et al. Safety of the 2011-12, 2012-13, and 2013-14 seasonal influenza vaccines in pregnancy: preterm delivery and specific malformations, a study from the case-control arm of VAMPSS. *Vaccine.* 2016;34(37):4450-4459.

24.	Lylianou D, Phousavath S, Vongphrachanh P, et al. Adverse events following immunization from pandemic influenza A (H1N1)-Laos 2010. *International Journal of Infectious Diseases.* 2012;16:e308-e309.

25.	McHugh L, Andrews RM, Lambert SB, et al. Birth outcomes for Australian mother-infant pairs who received an influenza vaccine during pregnancy, 2012-2014: the FluMum study. *Vaccine.* 2017;35(10):1403-1409.

26.	McHugh L, Binks MJ, Gao Y, et al. Influenza vaccination in pregnancy among a group of remote dwelling Aboriginal and Torres Strait Islander mothers in the Northern Territory: The 1+1 Healthy Start to Life study. *Communicable diseases intelligence (2018).* 2019;43.

27.	Mohammed H, Roberts CT, Grzeskowiak LE, Giles LC, Dekker GA, Marshall HS. Safety and protective effects of maternal influenza vaccination on pregnancy and birth outcomes: A prospective cohort study. *EClinicalMedicine.* 2020;26.

28.	Moro PL, Cragan J, Lewis P, Sukumaran L. Major Birth Defects after Vaccination Reported to the Vaccine Adverse Event Reporting System (VAERS), 1990 to 2014. *Birth Defects Research.* 2017;109(13):1057-1062.

29.	Moro PL, Marquez P. Reports of cell-based influenza vaccine administered during pregnancy in the Vaccine Adverse Event Reporting System (VAERS), 2013–2020. *Vaccine.* 2020.

30.	Nordin JD, Kharbanda EO, Benitez GV, et al. Maternal safety of trivalent inactivated influenza vaccine in pregnant women. *Obstetrics & Gynecology (New York).* 2013;121(3):519-525.

31.	Ohfuji S, Deguchi M, Tachibana D, et al. Safety of influenza vaccination on adverse birth outcomes among pregnant women: A prospective cohort study in Japan. *International journal of infectious diseases : IJID : official publication of the International Society for Infectious Diseases.* 2020;93:68-76.

32.	Omer SB, Goodman D, Steinhoff MC, et al. Maternal influenza immunization and reduced likelihood of prematurity and small for gestational age births: a retrospective cohort study. *PLoS Medicine.* 2011;8(5):e1000441.

33.	Peppa M, Thomas SL, Minassian C, et al. Seasonal influenza vaccination during pregnancy and the risk of major congenital malformations in live-born infants: A 2010-2016 historical cohort study. *Clinical infectious diseases : an official publication of the Infectious Diseases Society of America.* 2020.

34.	Phengxay M, Mirza SA, Reyburn R, et al. Introducing seasonal influenza vaccine in low-income countries: an adverse events following immunization survey in the Lao People's Democratic Republic. *Influenza and other respiratory viruses.* 2015;9(2):94‐98.

35.	Regan AK, Blyth CC, Mak DB, Richmond PC, Effler PV. Using SMS to monitor adverse events following trivalent influenza vaccination in pregnant women. *Australian and New Zealand Journal of Obstetrics and Gynaecology.* 2014;54(6):522-528.

36.	Regan AK, Tracey LE, Blyth CC, Richmond PC, Effler PV. A prospective cohort study assessing the reactogenicity of pertussis and influenza vaccines administered during pregnancy. *Vaccine.* 2016;34(20):2299-2304.

37.	Regan AK, Moore HC, Sullivan SG. Does influenza vaccination during early pregnancy really increase the risk of miscarriage? *Vaccine.* 2018;36(17):2227-2228.

38.	Richner JM, Jagger BW, Shan C, et al. Vaccine mediated protection against Zika virus-induced congenital disease. *Cell (Cambridge).* 2017;170(2):273-e212.

39.	Shatla MM, Khayat ME, Ahmed MM, et al. Effect of maternal antenatal influenza vaccination on adverse neonatal outcomes in terms of premature birth, small-for-gestational age and low birth weight: a comparative study. *International Journal of Medical Science and Public Health.* 2016;5(11):2378-2384.

40.	Vazquez-Benitez G, Kharbanda EO, Naleway AL, et al. Risk of preterm or small-for-gestational-age birth after influenza vaccination during pregnancy: caveats when conducting retrospective observational studies. *American Journal of Epidemiology.* 2016;184(3):176-186.

41.	Walsh LK, Donelle J, Dodds L, et al. Health outcomes of young children born to mothers who received 2009 pandemic H1N1 influenza vaccination during pregnancy: retrospective cohort study. *BMJ (Clinical research ed).* 2019;366:l4151.

42.	Wijnans L, Dodd CN, Weibel D, Sturkenboom M. Bell's palsy and influenza(H1N1)pdm09 containing vaccines: a self-controlled case series. *PLoS ONE.* 2017;12(5):e0175539.
